# Rotavirus disease and health care utilisation among children under 5 years of age in highly developed countries: a systematic review and meta-analysis

**DOI:** 10.1101/2020.12.03.20243071

**Authors:** Cristina Ardura-Garcia, Christian Kreis, Milenko Rakic, Manon Jaboyedoff, Maria Christina Mallet, Nicola Low, Claudia E. Kuehni

**Affiliations:** Institute of Social and Preventive Medicine, University of Bern, Bern, Switzerland; Service of Paediatrics, Department Women-Mother-Child, Lausanne University Hospital and University of Lausanne, Lausanne, Switzerland; Graduate School for Health Sciences, University of Bern, Bern, Switzerland; Paediatric Respiratory Medicine, Children’s University Hospital of Bern, University of Bern, Bern, Switzerland

**Author notes:** **Corresponding author:** Claudia E Kuehni. Mittelstrasse 43, 3012, Bern, Switzerland. Contributed equally (Cristina Ardura-Garcia, Christian Kreis).

**Keywords:** rotavirus, gastroenteritis, health care use, mortality, systematic review

## Abstract

**Background:** Rotavirus (RV) infection is the leading cause of diarrhoea-associated morbidity and mortality globally among children under 5 years of age. RV vaccination is available, but has not been implemented in many national immunisation plans, especially in highly developed countries. This systematic review aimed to estimate the prevalence and incidence of health care use for RV gastroenteritis (RVGE) among children aged under 5 years in highly developed countries without routine RV vaccination.

**Methods:** We searched MEDLINE and Embase databases from January 1^st^ 2000 to December 17^th^ 2018 for publications reporting on incidence or prevalence of RVGE-related health care use in children below 5 years of age: primary care and emergency department (ED) visits, hospitalisations, nosocomial infections and deaths. We included only studies with laboratory-confirmed RV infection, undertaken in highly developed countries with no RV routine vaccination plans. We used random effects meta-analysis to generate summary estimates with 95% confidence intervals (CI) and prediction intervals.

**Results:** We screened 4033 abstracts and included 74 studies from 21 countries. Average incidence rates of RVGE per 100 000 person-years were: 2484 (95% CI 697-5366) primary care visits, 1890 (1597-2207) ED visits, 500 (422-584) hospitalisations, 34 (20-51) nosocomial infections and 0.04 (0.02-0.07) deaths. Average proportions of cases of acute gastroenteritis caused by RV were: 21% (95% CI 16-26%) for primary care visits; 32% (25-38%) for ED visits; 41% (36-47%) for hospitalisations, 29% (25-34%) for nosocomial infections and 12% (8-18%) for deaths. Results varied widely between and within countries, and heterogeneity was high (I^2^>90%) in most models.

**Conclusion:** RV in children under 5 years causes many healthcare visits and hospitalisations, with low mortality, in highly developed countries without routine RV vaccination. The health care use estimates for RVGE obtained by this study can be used to model RV vaccine cost-effectiveness in highly developed countries.

**Take home message:** RV-caused illness leads to a high burden of health care usage in highly developed countries who have not introduced RV vaccination.

## Introduction

Rotavirus (RV) infection is the leading cause of diarrhoea-associated morbidity and mortality globally among children under 5 years of age [1]. RV-associated deaths are rare in high-income countries, but RV was estimated to account for 4.9 million episodes of diarrhoea among children younger than 5 years in Western Europe in 2016 [1]. The proportion of hospitalisations for acute gastroenteritis in children under 5 years caused by RV varies worldwide but has been reported to be 30-40% in both high- and low-income countries [2]. RV gastroenteritis (RVGE) therefore still poses a considerable burden on health care resources, although it is a vaccine-preventable disease.

Oral RV vaccines have been available since 2006 and have proven clinical efficacy against severe RVGE in different regions of the world, as summarised in a Cochrane review of randomized controlled trials in 2019 [3]. The World Health Organization (WHO) recommended the inclusion of RV vaccine in all national immunisation programmes in 2009, especially for countries at high risk of severe disease and mortality [4]. Most of the 98 countries that had included RV vaccination in their national immunisation plans in 2018 were low- and middle-income countries, which receive financial support from the global Vaccine Alliance (Gavi) [5]. In Europe, however, only one third of countries had introduced RV vaccination in their national vaccination programmes in 2016 [6]. The main barrier to the introduction of the RV vaccine in high-income countries is the perceived low burden of the disease, owing to low mortality rates [7,8]. High and middle income-countries also pay a higher price for the RV vaccine than countries supported by Gavi, because of a policy called “tiered pricing”. The low mortality and relatively high costs of the RV vaccine have led public health authorities in high-income countries to question the cost-effectiveness of the vaccine. However, the burden on healthcare of RV-related disease includes the morbidity that leads to primary care consultations, emergency care, hospitalisations and nosocomial infections, as well as mortality (Figure 1).

**Figure 1.**
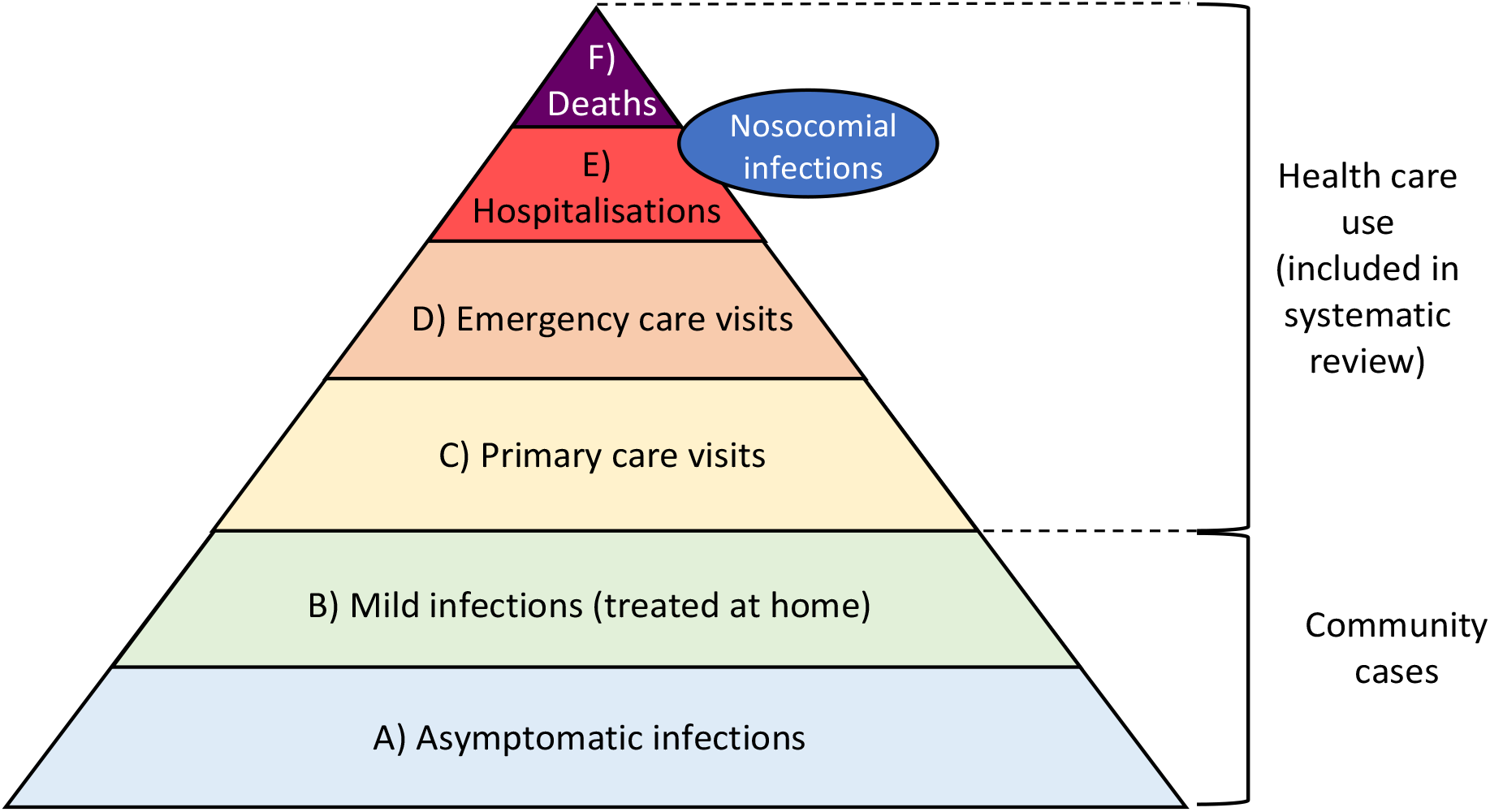
Contributions to rotavirus burden of health care use. Levels C through F were estimated in our study.

Previous systematic reviews have described the burden of RV-related health care use [2,9-15]. Most included studies from low and middle-income countries [2,11,12,14], where health care systems and gastroenteritis prevention strategies such as hand hygiene and adequate sanitation differ greatly from high-income countries. Most reviews did not include RV-related disease at all health care levels, from primary care visits to mortality [2,9-12,14]. Finally, the most recent reviews of RV health care use also include countries that have introduced routine RV vaccination [16]. To assess the cost-effectiveness of RV vaccines in high-income countries, we need RV-related health care use estimates that include all levels of care in these regions of the world. We conducted a systematic review and meta-analysis to estimate the incidence and prevalence of primary and emergency care visits, hospitalisations, nosocomial infections and deaths for RVGE among children under 5 years in highly developed countries that have not introduced routine RV vaccination.

## Methods

The protocol for this systematic review has been registered in the PROSPERO repository (CRD42019118069). We used the Preferred Reporting Items for Systematic Reviews and Meta-Analyses statement (PRISMA, research checklist online) [17] to report our findings.

### Eligibility criteria

We searched for scientific manuscripts reporting on health care use for RVGE in children under 5 years old in highly developed countries, before routine RV vaccination introduction. To obtain accurate estimates, we included only studies that used specific and reliable tests to diagnose RVGE and specified age ranges. We included studies that fulfilled all of the following criteria: (1) included children aged between 6 months or below and 47 months or more (up to 60 months) or between 0 and 24 months; (2) reported separate data for primary or emergency care visits, hospitalisations, nosocomial infections or deaths; (3) reported laboratory tests of stool samples positive for RV pathogen or antigens; and (4) were conducted in an eligible highly developed country. We defined highly developed countries as those with a good healthcare system (WHO mortality strata A: low adult mortality, very low child mortality [18]), a democratic government and a high-income economy (Organization for Economic Cooperation and Development (OECD) member [19])). The list of eligible countries is shown in Supplementary Table 1.

To reduce the risk of biases from studies with small sample sizes, misclassification of diagnosis and limited study period, we defined exclusion criteria based on sample size, year of publication and study duration. Specific exclusion criteria were: (1) use of only ICD codes in hospital discharge records or insurance billing data to classify RVGE cases, instead of laboratory-confirmed diagnosis; (2) used secondary data on RVGE in the analyses or data already published in other manuscripts; (3) were published before the year 2000; (4) were conducted after the introduction of routine or national rotavirus vaccination (more than 80% coverage) and did not report separate results for a sub-period preceding general vaccination or a trial arm of unvaccinated children; (5) had a study population of fewer than 500 children (except for studies analysing the rare outcomes of nosocomial infection or mortality); or (6) had a study period of less than 12 months (to exclude reports of healthcare use only during RV peak seasons).

### Information sources and search strategy

We searched the Embase and MEDLINE databases on December 17, 2018. Our search strings included medical subject headings (MeSH) or thesaurus terms and free text key words pertaining to (1) rotavirus, (2) disease outbreak, surveillance or epidemiology, (3) names of major geographic regions or cities in eligible countries and (4) infancy and childhood. The complete search strings are presented in the supplementary material. We used an automated procedure to eliminate duplicates and generated an EndNoteX8 (Clarivate Analytics, Philadelphia, Pennsylvania, United States of America, USA) library to screen retrieved records.

### Study selection

We screened titles and abstracts to assess eligibility using an accelerated process (screening by one reviewer, followed by confirmation of records for exclusion by a second reviewer) [20]. Two independent reviewers then screened the full-text manuscripts of potentially eligible studies in duplicate. Disagreements were settled by consensus or by discussion with a third researcher. We recorded the reasons for exclusion of full-text manuscripts in an Excel spreadsheet.

### Data extraction

We extracted the data from all eligible publications into a pre-piloted standard data extraction form into a secure online database (Research Electronic Data Capture, REDCap, Vanderbilt University, Tennessee, USA). Data extraction was done by one reviewer and checked by a second reviewer (e.g. missing details or to confirm important data). We extracted information on the publication, study characteristics and the number and characteristics of the participants, the authors’ definitions of the outcome measure and the specific laboratory procedure for identification of causal pathogen. For each outcome (primary care visit, emergency care visit, hospitalisation, nosocomial infection or death), we extracted numerical results (absolute number, proportion, rate) with 95% CI, if available. We extracted the total number of children with acute gastroenteritis, the number of stool samples tested, the number of positive RV stools samples and the person-years of the population at risk, for each outcome. A detailed list of items extracted is described in the online supplementary material.

### Risk of bias assessment

We assessed the risk of bias in individual studies by adapting the tool proposed by Hoy et al. [21]. This tool was specifically designed to assess population-based prevalence studies of back pain. The checklist contains 10 items to assess external and internal validity of studies which are classified as: Yes (low risk) or No (high risk). Questions 1-4 assess external validity: study population representativeness of national and target populations, selection methods and non-response bias (which we defined as less than 75% of children with acute gastroenteritis tested for RV, or no clear report of this proportion). Questions 5-10 assess internal validity: outcome data collection, definition of outcomes, reliability of outcome measures, length of study period and the use of appropriate numerators and denominators.

### Synthesis of results and analysis

For each study and outcome, we used the number of positive RV stools samples and the person-years of the population at risk to calculate incidence rates, and the number of cases of acute gastroenteritis and the number of RV positive stool samples to calculate pooled proportions. For incidence, if the number of RVGE cases were not reported, we calculated it based on person-years and reported incidence rates. Similarly, if the number of person-years was not reported, we calculated it based on the incidence rate and 95% confidence interval if reported. For proportions, if only the numerator (RV positive stool samples) or the denominator (number of all cases of acute gastroenteritis) were reported, we used the reported proportion of RVGE to calculate the other number. Nosocomial infections were commonly reported per number of hospitalisations or number of hospital-days. If studies reported nosocomial infections occurring both during hospitalisation and after hospital discharge separately, we only used the in-hospital diagnoses.

We expected variability in health care use for RVGE between countries due to differences in climate and health care systems. We therefore performed random effects meta-analysis to calculate summary estimates of average incidence rates or proportions (with 95% CI). We used the Freeman-Tukey Double arcsine transformation to calculate the overall incidence rate or proportion [22]. We examined heterogeneity graphically, using forest plots, statistically by calculating the I^2^ and Tau-squared statistics [23], and in subgroup analyses at the country level. We calculated the prediction interval to indicate the range of the predicted proportion or incidence rate in a new study conditional on the previous studies [23] [24]. However, if the heterogeneity between studies was so large as to render the prediction interval uninformative, the prediction interval was not plotted along with the forest plots. We did not test the included studies for potential risk of publication bias because the relevance for studies of prevalence or incidence is uncertain. We used the R language for statistical computing for all analysis, using the metaprop and metarate functions of the package ‘meta’ [25] and package ‘epitools’ [26].

We performed two sensitivity analyses to explore possible causes of heterogeneity. First, we assessed incidence rates and proportions in studies that included only children under 2 years of age, as severe RVGE affects particularly children of this age range. For this, we included all studies that included children aged between 0 and 24 months. In a second sensitivity analysis we assessed if the risk of bias in the studies affected the pooled estimates for children under 5 years old. For this analysis, we only included studies that had a low risk of bias score for selection and non-response bias, i.e. items 3 and 4 on the Hoy et al. risk of bias tool [21].

## Results

Our electronic search identified 4033 records after removing duplicates, of which we selected 1336 based on screening of titles and abstracts. Of these, we included 77 full-text manuscripts reporting on 74 studies in the systematic review [27-101] (Figure 2). The included studies were conducted in 21 different countries in North America, Europe, Asia and Oceania.

**Figure 2.**
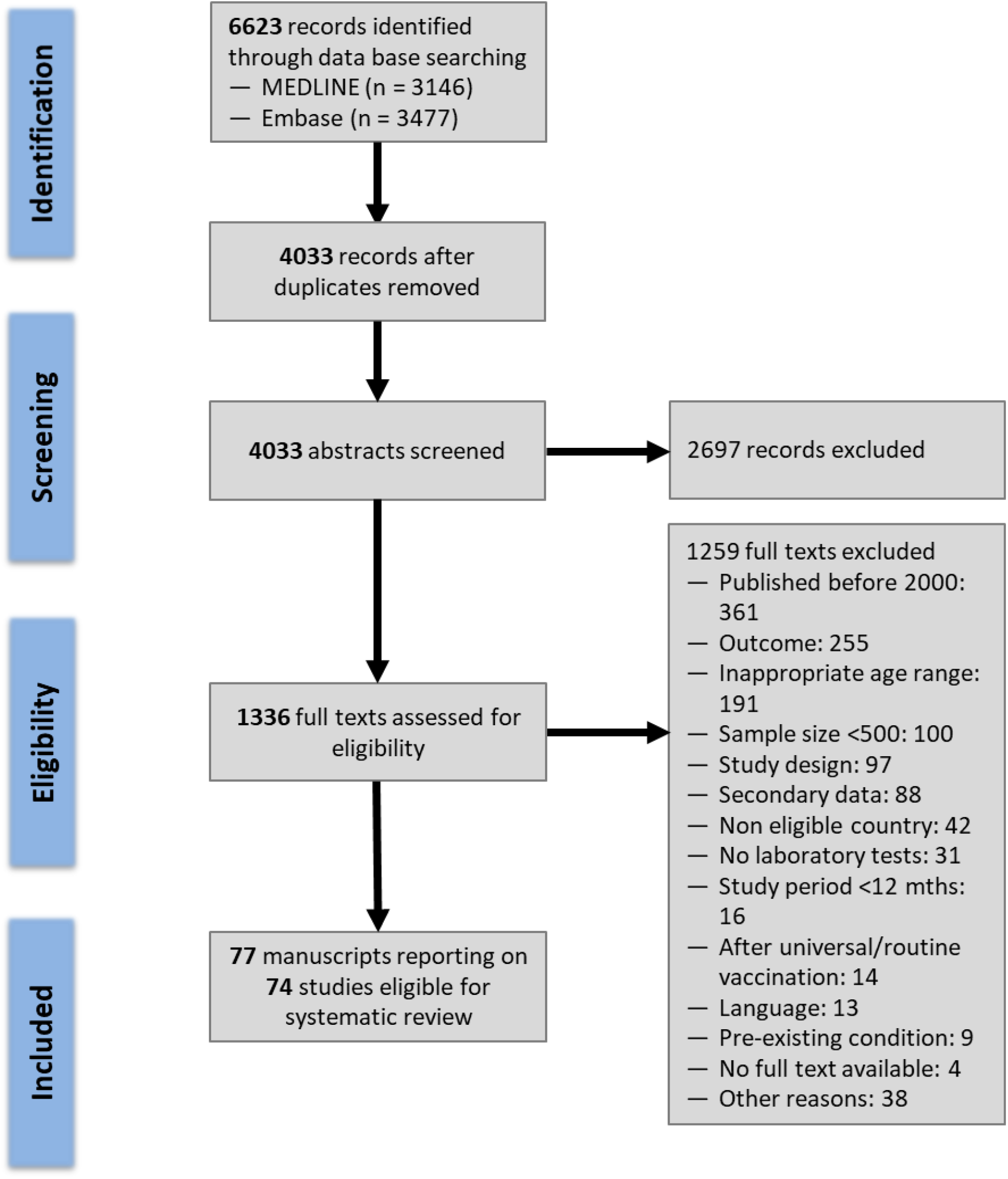
Rotavirus burden of health care use: flow diagram of study selection.

We now describe results for different levels of health care use: primary care, emergency care, hospitalisations, nosocomial infections and deaths. For each level of health care use, we present summary estimates of incidence rates and proportions of acute gastroenteritis caused by RV infection in children under 5 years of age (Table 1).

**Table 1.** Summary table with pooled estimates of health care use for rotavirus gastroenteritis in children in highly developed countries with no routine rotavirus vaccination programmes.

|  | **Outcome measure,**<br>**number of studies (n)** | **Under 5 years old** | **Under 2 years old** | **Under 5 years, low**<br>**risk of bias#** |
| --- | --- | --- | --- | --- |
| **Primary care visits** | Proportion (95% CI) | 21% (16-26%), 14 | 24% (14-35%), 2 | 17% (10-26%), 7 |
|  | Incidence per 100 000 person-years (95% CI) | 2484 (697-5366), 3 | 4390 (2916-6161), 2 | - |
| **Emergency care visits** | Proportion (95% CI) | 32% (25-38%), 19 | 34% (9-66), 2 | 29% (20-39%), 11 |
|  | Incidence per 100 000 person-years (95% CI) | 1890 (1597-2207), 1 | 1205 (193-3072), 2 | - |
| **Hospitalisations** | Proportion (95% CI) | 41% (36-47%), 45 | 50% (32-68%), 8 | 40% (32-47%), 30 |
|  | Incidence per 100 000 person-years (95% CI) | 500 (422-584), 13 | 826 (650-1024), 9 | 544 (470-623), 10 |
| **Nosocomial infections** | Proportion (95% CI) | 29% (25-34%), 3 | 53% (46-60%), 3 | 29% (25-34%), 3 |
|  | Incidence per 100 000 person-years (95% CI) | 34 (20-51), 3 | 95 (90-100), 1 | 43 (36-50), 2 |
|  | Incidence per 1000 hospital-days (95% CI) | 0.63 (41-90), 3 | 7.09 (1-17), 4 | 0.61 (0.33-0.98), 2 |
|  | Incidence per 1000 hospitalisations (95% CI) | 1.5 (1-2.1), 1 | 18 (3-46), 3 | - |
| **Mortality** | Proportion (95% CI) | 12% (8-18%), 1 | - | - |
|  | Rate per 100 000 person-years (95% CI) | 0.04 (0.02-0.07), 2 | - | 0.04 (0.02-0.07), 2 |
|  | Rate per 100 000 hospitalisations (95% CI) | 129 (81-187), 1 | - | - |

#: Pooled estimates including only studies with low risk of bias score for selection and non-response bias, i.e. items 3 and 4 on the Hoy et al. risk of bias tool \*Results from one study alone (not a pooled estimate).

### Primary care visits

The incidence rate of primary care visits for RVGE was reported by three studies (Figure 3); 1036 and 2270 per 100 000 children per year in Italy [75] and 4841 in Germany [77]. The pooled incidence rate was 2484 (95%CI: 697-5366) per 100 000 person-years (I^2^: 99%). The summary estimate of the proportion of primary care visits for acute gastroenteritis caused by RV infection was 21% (95% CI: 16-26%; prediction interval: 5-44%; I^2^: 98%) in 14 studies from 9 countries (Figure 4). The results varied between countries, with the highest proportion found in a study from the USA (45%, 95% CI: 40-49%) [50] and the lowest in studies from Spain (9%, 95% CI: 4-15%) [74,86].

**Figure 3.**
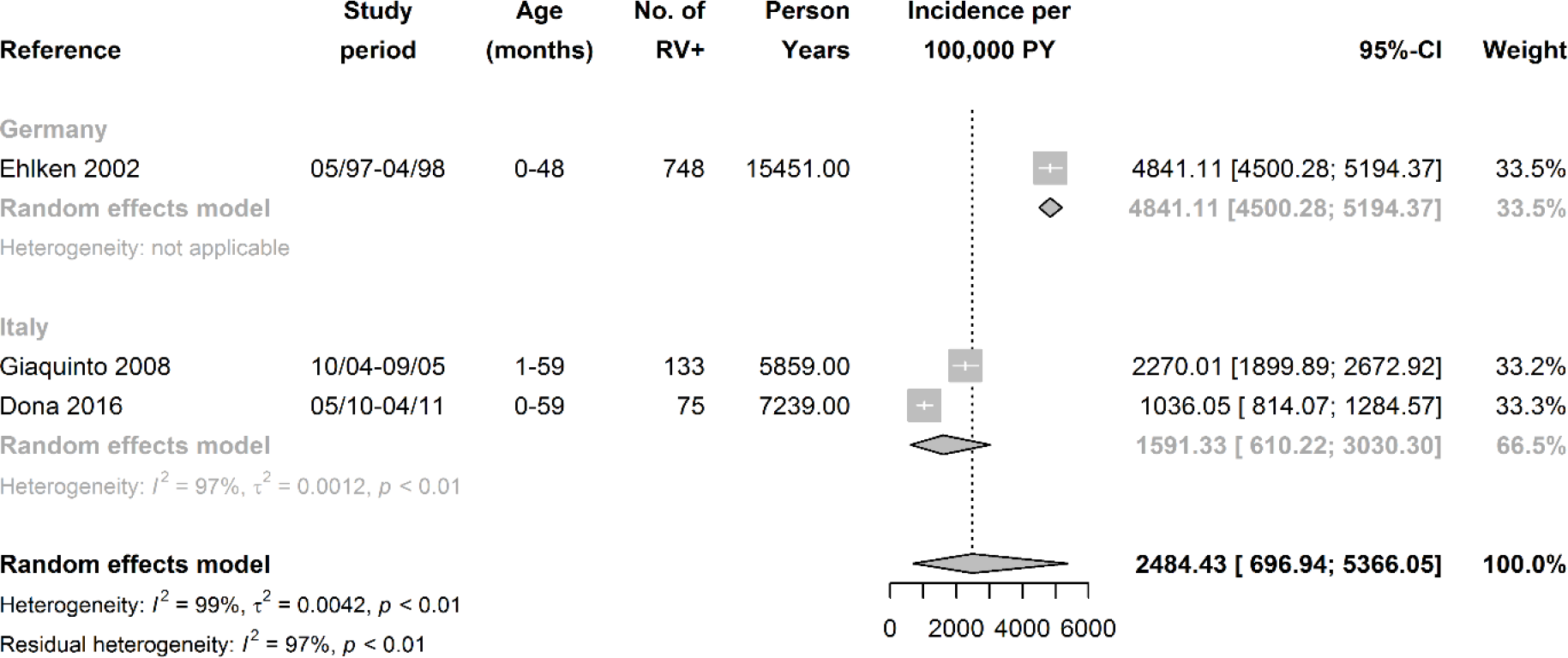
Incidence rate of primary care visits for RVGE of children aged 0-5 years per 100,000 person-years (forest plot) RVGE: Rotavirus gastroenteritis; RV+: stools samples that tested positive for rotavirus; PY: person-years; CI: Confidence Interval

**Figure 4.**
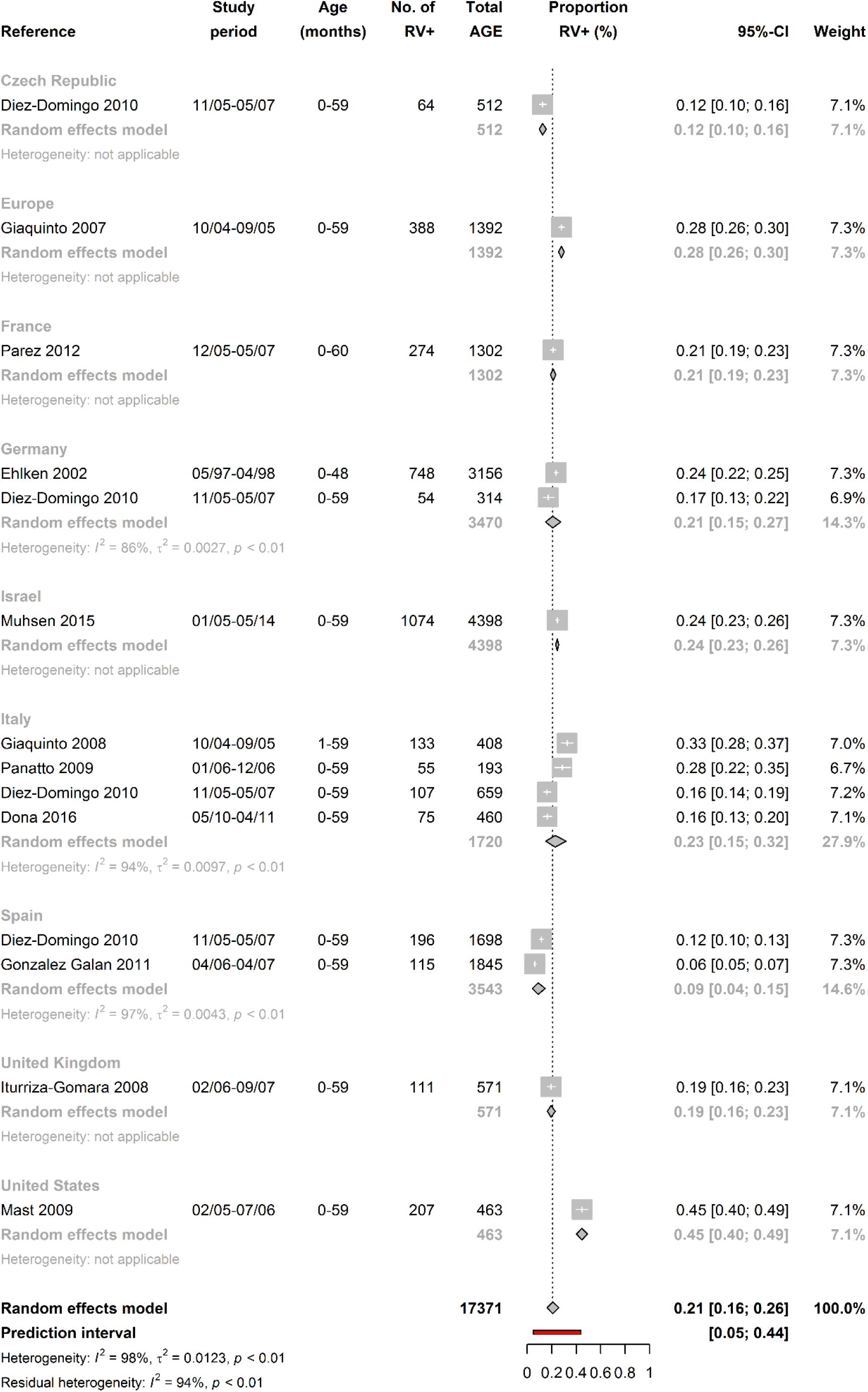
Proportion of primary care visits for RVGE among all visits for acute gastroenteritis of children aged 0-5 years (forest plot) RVGE: Rotavirus gastroenteritis; RV+: stools samples that tested positive for rotavirus; AGE: acute gastroenteritis; CI: Confidence Interval

### Emergency care visits

The incidence rate of emergency care visits for RVGE was reported by only one study from Italy, with an incidence rate of 1890 per 100 000 children per year (95% CI: 1597-2207) [83]. The summary estimate of the proportion of emergency care visits for acute gastroenteritis caused by RV infection was 32% (95% CI: 25-38%; prediction interval: 6-65%; I^2^: 98%) in 19 studies from 11 countries (Figure 5). The highest proportion was found in a study performed in multiple European countries (Rotavirus Gastroenteritis Epidemiology and Viral Types in Europe Accounting for Losses in Public Health and Society, REVEAL, study: Belgium, France, Germany, Italy, Spain, Sweden and the United Kingdom, UK) (49%, 95% CI: 46-53%) [84] and the lowest in a study from Greece (11%, 95% CI: 5-18%) [95].

**Figure 5.**
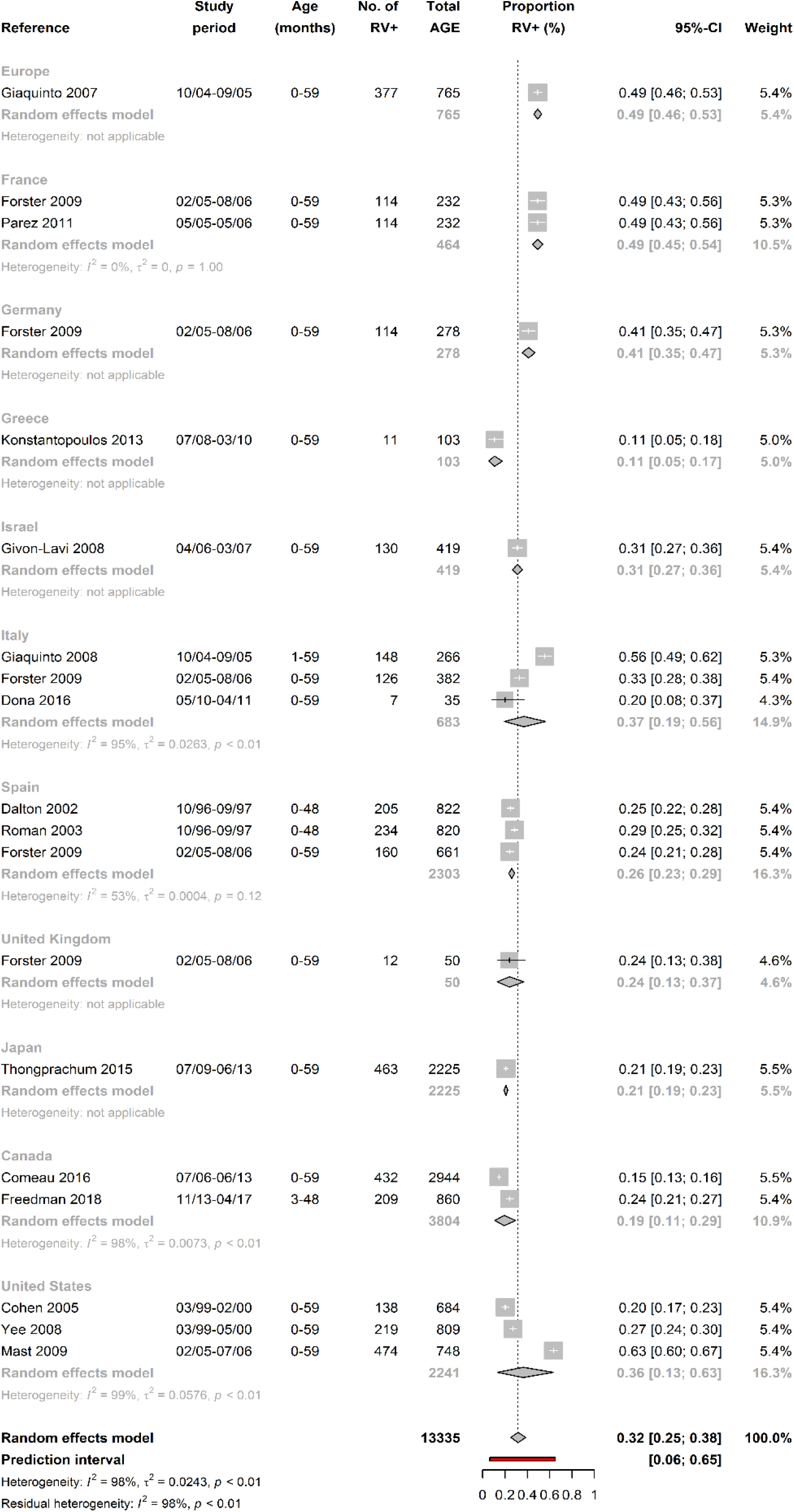
Proportion of emergency department consultations for RVGE among all consultations for acute gastroenteritis of children aged 0-5 years (forest plot) RVGE: Rotavirus gastroenteritis; RV+: stools samples that tested positive for rotavirus; AGE: acute gastroente3rit1is;

### Hospitalisations

The incidence rate of hospitalisations for RVGE was reported by 13 studies from 9 countries (Figure 6) and ranged from 149 (Italy) [83] to 766 (Austria) [44] per 100 000 person-years. The summary incidence rate was 500 (95%CI: 422-584; prediction interval 221-892; I^2^: 100%) per 100 000 children per year. The summary estimate of the proportion of hospitalisations for acute gastroenteritis caused by RV infection was 41% (95% CI: 36-47%; prediction interval: 9-79%; I^2^: 100%) in 45 studies from 16 countries (Figure 7). The highest proportions were reported in studies from France (64%, 95% CI: 59-70%) [56,80] and the lowest in studies from Greece (19%, 95% CI: 16-23%) [47,54,95]. Summary estimates by country are shown in Supplementary Figure 1.

**Figure 6.**
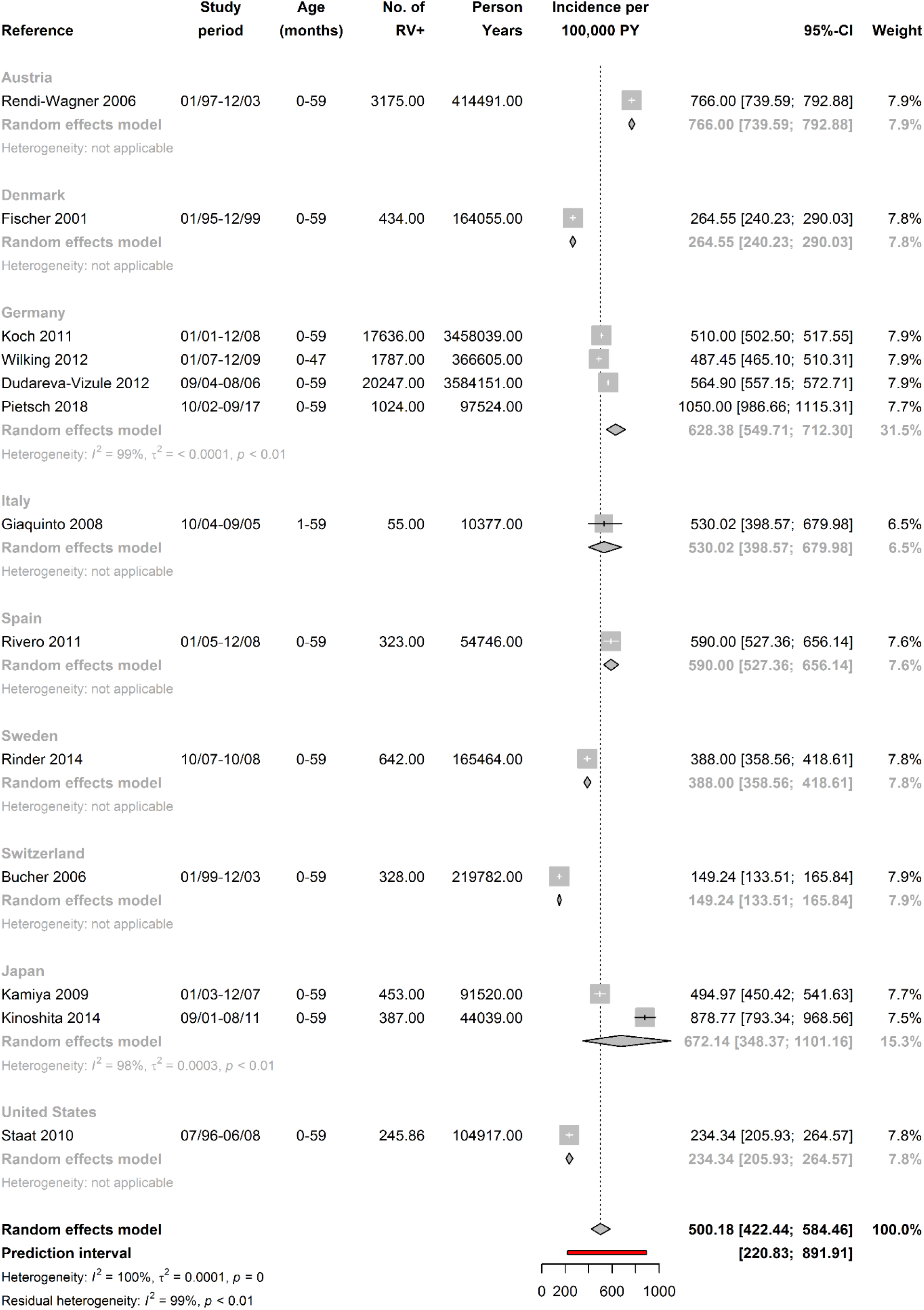
Incidence rate of hospitalisations for RVGE of children aged 0-5 years per 100,000 person-years (forest plot) RVGE: Rotavirus gastroenteritis; RV+: stools samples that tested positive for rotavirus; PY: person-years; CI: Confidence Interval

**Figure 7.**
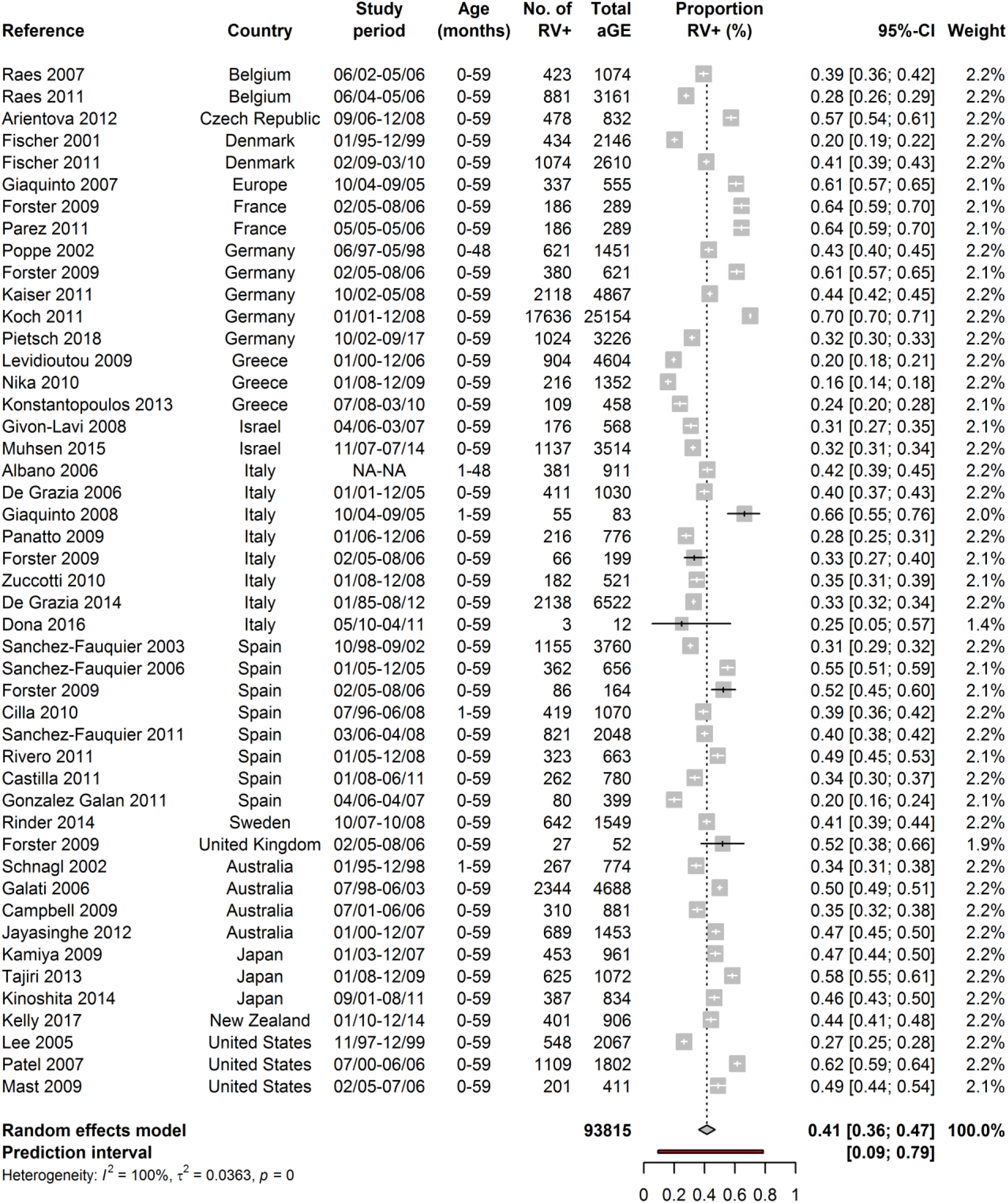
Proportion of hospitalisations for RVGE among all hospitalisations for acute gastroenteritis of children aged 0-5 years (forest plot without country estimates) RVGE: Rotavirus gastroenteritis; RV+: stools samples that tested positive for rotavirus; AGE: acute gastroenteritis; CI: Confidence Interval

### Nosocomial infections

The incidence rate of RVGE nosocomial infections per 100 000 person-years was reported by 3 studies from 3 countries [65,76,95] (Supplementary Figure 2). The summary incidence rate was 34 (95% CI: 20-51) per 100 000 children under 5 years old per year in the population, with high heterogeneity (I^2^: 91%).The incidence rate of RVGE nosocomial infections per 1000 hospital-days was reported by 3 other studies from 3 countries [32,56,80], with a summary incidence rate of 0.63 cases (95% CI: 0.41-0.90; prediction interval: 0-7.3; I^2^: 91%) per 1000 hospital-days (Supplementary Figure 3). The incidence proportion of RVGE nosocomial infections per 1000 hospitalisations was reported by one study from New Zealand as 1.5 (95% CI: 0.96-2.15) [92]. The proportion of nosocomial infections of acute gastroenteritis caused by RV in children under 5 years old hospitalised for reasons other than acute gastroenteritis was reported by 3 studies from 3 countries [79,82,95] (Supplementary Figure 4). The pooled proportion was 29% (95% CI: 25-34%; prediction interval: 2-72%; I^2^: 31%).

### Mortality

The mortality rate for RVGE in children under 5 years old was reported by 2 studies from Germany, with a pooled estimate of 0.04 deaths (95% CI: 0.02-0.07; I^2^: 0%) per 100 000 person-years [36,94] (Figure 8). The proportion of deaths for acute gastroenteritis caused by RV was reported by one study from the UK as 12% (95% CI: 8-18%) [89]. The same study also reported mortality for RVGE as 129 (95% CI 81-187) deaths per 100 000 hospitalisations in children under 5 years.

**Figure 8.**
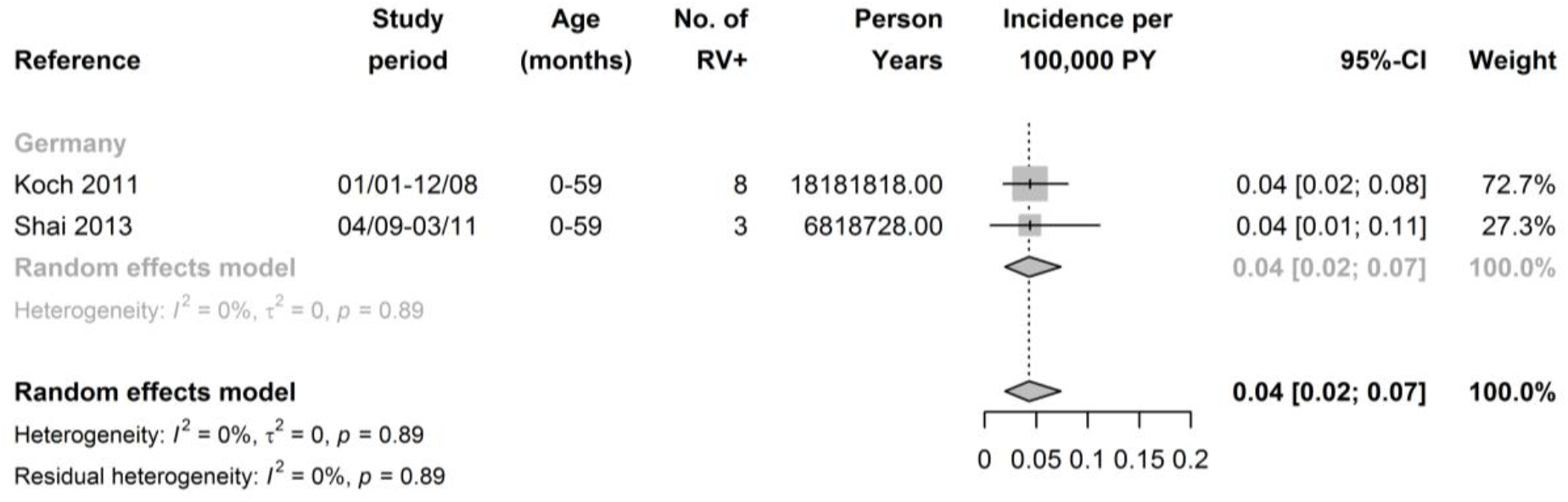
Mortality rate due to RVGE of children aged 0-5 years per 100,000 person-years (forest plot) RVGE: Rotavirus gastroenteritis; RV+: stools samples that tested positive for rotavirus; PY: person-years; CI: Confidence Interval

### Risk of bias

When assessing external validity (Supplementary Figure 5), 41 (55%) studies had a study population that was a good representation of the national population (question 1); the rest of the studies included only one centre or several centres but not from different regions in the country. In 55 (74%) studies, all children with acute gastroenteritis seen during the study period in the study population were included, or the studies tested a random sample of children for RV. The remaining studies included only those that agreed to participate or the reason for testing for RV was unclear or not reported (question 3). In 54 (73%) studies, the risk of non-response bias was low. When assessing internal validity, 58 (78%) studies used the same method to test all the stool samples for RV. One fifth of the studies (17 of 74) fulfilled all 10 items, while 23 (31%) fulfilled all the external validity items and 50 (68%) fulfilled all the internal validity items.

### Sensitivity analyses

We selected studies including children 0-24 months old for the first sensitivity analysis (Supplementary Figures 6-14). The incidence rates of RVGE were higher among children under 2 years of age for primary care visits, hospitalisations and nosocomial infections, but not for emergency care visits, when compared to the total population of under 5-year-olds. There were no studies reporting mortality rates due to RVGE in children under 2 years of age. The summary proportions of primary and emergency care visits for gastroenteritis caused by RV were slightly higher than those from children aged under 5 years (24% for primary care compared to 21% in under 5s, and 34% for emergency care compared to 32% in under-5s), whereas the summary proportions of hospitalisations reached 50% (compared to 41% in under-5s) and of nosocomial infections 53% (compared to 29% in under-5s).

We included only studies with low risk of bias for the second sensitivity analysis (Supplementary Figures 15-21). Overall, the pooled estimates of the incidence rates and proportions of the five different health care outcomes changed little when we excluded studies with lower external validity. The proportion of RVGE-related visits to primary and emergency care, and hospitalisations were slightly lower, whereas the estimates of the incidence of hospitalisations and RVGE nosocomial infections per 100 000 person-years were higher than in the analyses including all studies. The two studies included in the pooled analysis for mortality rate per 100 000 children under 5 years of age in the main analysis had low risk of bias (Figure 8).

## Discussion

This systematic review and meta-analysis found that, in highly developed countries with no RV routine vaccination, every year an average of 500 children (95% CI 422-584) are hospitalised for RVGE per 100 000 children under 5 years. Emergency care visits and primary care visits occurred 4 to 5 times more often. Mortality for RVGE was very low (0.04 deaths per 100 000 person-years; 95% CI: 0.02-0.07). In higher levels of the health care use pyramid (Figure 1), the summary estimate of the proportion of acute gastroenteritis caused by RV was higher (21% for primary care visits, 40% for hospitalisations). For nosocomial acute gastroenteritis, an average of 29% (25-34%) were caused by RVGE. In children under 2 years old, the incidence rate of primary care visits, hospitalisations and nosocomial infections for RVGE were much higher than those found in children under 5 years old, while the proportions of RVGE among all acute gastroenteritis, did not vary greatly. There were large variations between and within countries, and heterogeneity was high (>90%) in most models.

### Strengths and limitations

This systematic review summarised both incidence rates and proportions of RVGE disease across all levels of health care use, from primary care to hospitalisations, including nosocomial infections and mortality. This allowed to put together a complete picture of the burden of RVGE in highly developed countries with no routine RV vaccination programmes. We applied inclusion criteria that attempted to reduce small study and measurement biases. We did sensitivity analyses to assess possible causes of heterogeneity. Age distribution in study population is a possible cause of heterogeneity as younger children had higher incidence rates of health care use for RVGE. Studies with low risk of bias showed similar pooled estimates to those of the main analysis, though there were no low risk of bias studies reporting on primary care and emergency care visits incidence rate. We therefore think that risk of bias is unlikely to explain the observed heterogeneity.

This review also has some limitations. First, most models showed very high heterogeneity, both between and within countries. This may be due to differences in health care systems or climate, or both, so the average summary estimates may not be applicable to any specific settings. Nevertheless, our results for hospitalisations, for example, are rather similar to previously published summary estimates from highly developed countries [14,15]. Second, some of the included studies had a high risk of bias, especially when assessing their external validity. However, the pooled estimates did not vary greatly when excluding the high risk of bias studies in the sensitivity analysis. Finally, the proportion of children that had received the RV vaccine may have varied from one study to another. Even though we excluded studies undertaken after RV vaccine introduction in national immunisation programmes, a study from 2001 would have no RV vaccinated children, while a study from 2010 may have had a 10% of RV vaccinated children (paid privately). We did extract these data from the studies when available, but this was not always described.

### Findings in relation to other studies

For every hospitalisation for RVGE, we estimated that there were an average of 4 emergency care visits and 5 primary care visits. The ratio between primary care visits and hospitalisations was similar when including only children under 2 years old. A 2017 European Centre for Disease Prevention and Control (ECDC) report on RV burden in the European Union also found that there are 2-4 times more visits to the emergency department or other outpatient facilities caused by RVGE than hospitalisations [15]. However, summary proportions of acute gastroenteritis caused by RV were twice as high for primary care than for hospitalisations (21% vs 40%). Indeed, RV may cause a more severe gastroenteritis with a higher degree of dehydration requiring hospital admission more often than other gastrointestinal pathogens. One previous meta-analysis reported on RVGE burden estimates for different levels of health care but included only studies from China. In this review, the proportion of RV disease among children under 5 years with acute gastroenteritis was 33% for outpatient visits and 43% for inpatients, very similar to our summary estimates for emergency care visits and hospitalisations [13].

We found differences between studies reporting only on children aged less than 2 years, and studies reporting on under 5-year-olds. The summary incidence rates of primary and emergency care visits and hospitalisations for RVGE in children under 2 years of age were 1.7 times higher than in children under 5. We also found a higher incidence of nosocomial RVGE among children under 2 years of age (more than 10 times that of children under 5 years). The higher incidence of RVGE in children under 2 years of age, especially severe RVGE requiring hospitalisations, has been reported [12] and is explained by the higher risk of dehydration at a younger age. This finding could account for part of the variation in incidence and proportions found between studies depending on the age distribution. We could not study this further because not all studies reported the age distribution of their study population. However, the incidence rate of RVGE emergency care visits for children aged under 2 years was lower than that for children aged under 5 years. This estimate was reported by only one small study with a high risk of bias and may not be as reliable as the one obtained for hospitalisations.

Hospitalisations for RVGE have been studied worldwide, and several systematic reviews and meta-analysis focusing on different areas of the world reported on this outcome. Two reviews that included both high and low-income countries found pooled incidence rates for RVGE hospitalisations similar to our high-income countries estimates. The ECDC report found 49 studies reporting on RVGE hospitalisations in countries from the European Union and the European Economic Area, irrespective of the level of development. The reported incidence ranged from 100-1190 per 100 000 person years (most of which were between 300-600) and the proportion of all acute gastroenteritis hospitalisations ranged between 26-69% [15], comparable to our findings. A worldwide review including 242 studies found also a very similar overall proportion of RV-attributable hospitalisations for acute gastroenteritis in children under 5 years old (38%; 95% CI: 36-40%) [12]. However, this proportion was much lower in studies seeking multiple pathogens than in those testing for RV alone (20% in studies with 5– 13 pathogens vs. 39% in single-pathogen studies, p<0.0001) [12]. This is another possible cause of heterogeneity that we could not study in our review as we did not collect this data. Other worldwide reviews have found that the incidence of RV hospitalisations does not vary much with the level of economic development (median RV admissions per 1000 children under 5 years per year: 2.0 vs. 1.9 in low- and high-income regions, respectively) [14].

### Implications for health care and future research

The results from this meta-analysis can be used to model the cost-effectiveness of RV vaccine introduction into national immunisation plans in highly developed countries. Economic evaluations will provide useful information for public health authorities when deciding upon immunisation strategies for RV. We observed a wide variation of estimates of RVGE health care use between and within countries in our review, presumably due to differences in health care systems, climate and prevalence of other gastrointestinal pathogens, but also to differences in study designs (use of hospital records vs. cohort studies with invited participants), specific locations (multicentre vs. single centre studies) and laboratory methods used. This has resulted in the high heterogeneity observed in our pooled models. Researchers assessing RV-related health care in future could obtain more robust results by using standardised prospective methods across different countries.

## Conclusion

RV causes a considerable number of health care visits and hospitalisations for gastroenteritis in highly developed countries without routine RV vaccination. The summary estimate of the proportion of acute gastroenteritis caused by RV was twice as high for hospitalisations (41%) as for primary care (21%), positioning RV as one of the main pathogens of severe gastroenteritis requiring hospitalisation in children under 5 years. Even though RVGE mortality is low in highly developed countries, this vaccine-preventable disease poses a considerable burden on health care systems.

## Data Availability

The data used for this manuscript was extracted from published papers.

## Role of the Funding Source

The study was partly supported by a research grant from GlaxoSmithKline Biologicals SA (GSK study identifier HO-18-19250). GlaxoSmithKline Biologicals SA participated in the study design but had no role in the data collection, analysis, interpretation of the results and writing of the manuscript. GlaxoSmithKline Biologicals SA was provided the opportunity to review a preliminary version of this manuscript for factual accuracy, but the authors are solely responsible for final content and interpretation.

## Authors contributions

Claudia E Kuehni, Nicola Low, Cristina Ardura-Garcia and Christian Kreis conceptualised and designed the study. Cristina Ardura-Garcia, Christian Kreis, Milenko Rakic, Manon Jaboyedoff and Christina Mallet performed the screening and data extraction. Christian Kreis analysed the data. Claudia E Kuehni, Nicola Low, Cristina Ardura-Garcia and Christian Kreis interpreted the results. Cristina Ardura-Garcia drafted the manuscript. All authors critically revised the manuscript and approved the final manuscript as submitted.

## Notes

### Author Declarations

This is a systematic review and meta-analysis. It is threfore exempt of ethics committee approvals.

