## Supplemental Material for "Rotavirus disease and health care utilisation among children under 5 years of age in highly developed countries: a systematic review and meta-analysis"

### Supplementary Online Material: Index


| Item | Page |
|---|---|
| Supplementary Table 1: List of eligible countries | 3 |
| Online supplementary methods: Detailed search string | 4 |
| Online supplementary methods: Detailed items extracted | 9 |
| Supplementary Figure 1. Proportion of hospitalisations for RVGE among all hospitalisations for acute gastroenteritis of children aged 0-5 years (forest plot with country estimates) | 10 |
| Supplementary Figure 2. Incidence rate of RVGE nosocomial infections among children aged 0-5 years hospitalised for reasons other than acute gastroenteritis per 100 000 person-years (forest plot) | 12 |
| Supplementary Figure 3. Incidence rate of RVGE nosocomial infections among children aged 0-5 years hospitalised for reasons other than acute gastroenteritis per 1000 hospital-days (forest plot) | 12 |
| Supplementary Figure 4. Proportion of acute gastroenteritis nosocomial infections caused by RV among children aged 0-5 years hospitalised for reasons other than acute gastroenteritis (forest plot) | 13 |
| Supplementary Figure 5. Number of studies that fulfilled each item of Hoy et al. risk of bias tool (N= 74) | 13 |
| Supplementary Figure 6. Incidence rate of primary care visits for RVGE of children aged 0-2 years per 100 000 person-years (forest plot) | 14 |
| Supplementary Figure 7. Proportion of primary care visits for RVGE among all visits for acute gastroenteritis of children aged 0-2 years (forest plot) | 14 |
| Supplementary Figure 8. Incidence rate of emergency department consultations for RVGE of children aged 0-2 years per 100 000 person-years (forest plot) | 15 |
| Supplementary Figure 9. Proportion of emergency department consultations for RVGE among all consultations for acute gastroenteritis of children aged 0-2 years (forest plot) | 15 |
| Supplementary Figure 10. Incidence rate of hospitalisations for RVGE of children aged 0-2 years per 100 000 person-years (forest plot) | 16 |
| Supplementary Figure 11. Proportion of hospitalisations for RVGE among all hospitalisations for acute gastroenteritis of children aged 0-2 years (forest plot) | 17 |
| Supplementary Figure 12. Incidence proportion of RVGE nosocomial infections among children aged 0-2 years hospitalised for reasons other than acute gastroenteritis per 1000 hospitalisations (forest plot) | 18 |
| Supplementary Figure 13. Incidence rate of RVGE nosocomial infections among children aged 0-2 years hospitalised for reasons other than acute gastroenteritis per 1000 hospital-days (forest plot) | 18 |
| Supplementary Figure 14. Proportion of acute gastroenteritis nosocomial infections caused by RV among children aged 0-2 years hospitalised for reasons other than acute gastroenteritis (forest plot) | 19 |
| Supplementary Figure 15. Proportion of primary care visits for RVGE among all visits for acute gastroenteritis of children aged 0-5 years, including only studies with good external validity (forest plot) | 20 |
| Supplementary Figure 16. Proportion of emergency department consultations for RVGE among all consultations for acute gastroenteritis of children aged 0-5 years, including only studies with good external validity (forest plot) | 21 |
| Supplementary Figure 17. Incidence rate of hospitalisations for RVGE of children aged 0-5 years per 100 000 person-years, including only studies with good external validity (forest plot | 22 |

| | |
|---|---|
| Supplementary Figure 18. Proportion of hospitalisations for RVGE among all hospitalisations for acute gastroenteritis of children aged 0-5 years, including only studies with good external validity (forest plot) | 23 |
| Supplementary Figure 19. Incidence rate of RVGE nosocomial infections among children aged 0-5 years hospitalised for reasons other than acute gastroenteritis per 100 000 person-years, including only studies with good external validity (forest plot) | 25 |
| Supplementary Figure 20. Incidence rate of RVGE nosocomial infections among children aged 0-5 years hospitalised for reasons other than acute gastroenteritis per 1000 hospital-days, including only studies with good external validity (forest plot) | 25 |
| Supplementary Figure 21. Proportion of acute gastroenteritis nosocomial infections caused by RV among children aged 0-5 years hospitalised for reasons other than acute gastroenteritis, including only studies with good external validity (forest plot) | 26 |

**Supplementary Table 1:** List of eligible countries

| World Region | WHO Mortality A | OECD Members |
|---|---|---|
| **European Region** | Austria | Austria |
| | Belgium | Belgium |
| | Czech Republic | Czech Republic |
| | Denmark | Denmark |
| | Finland | Finland |
| | France | France |
| | Germany | Germany |
| | Greece | Greece |
| | Iceland | Iceland |
| | Ireland | Ireland |
| | Israel | Israel |
| | Italy | Italy |
| | Luxembourg | Luxembourg |
| | Netherlands | Netherlands |
| | Norway | Norway |
| | Portugal | Portugal |
| | Slovenia | Slovenia |
| | Spain | Spain |
| | Sweden | Sweden |
| | Switzerland | Switzerland |
| | United Kingdom | United Kingdom |
| | *Andorra* | |
| | *Croatia* | |
| | *Malta* | |
| | *Monaco* | |
| | *San Marino* | |
| | | *Estonia* |
| | | *Hungary* |
| | | *Latvia* |
| | | *Lithuania* |
| | | *Poland* |
| | | *Slovak Republic* |
| | | *Turkey* |
| **Region of the Americas** | Canada | Canada |
| | United States of America | United States of America |
| | *Cuba* | |
| | | *Chile* |
| | | *Mexico* |
| **Asia and Western Pacific Region** | Australia | Australia |
| | Japan | Japan |
| | New Zealand | New Zealand |
| | *Brunei Darussalam* | |
| | *Singapore* | |
| | | *Korea* |

In italics and light grey: countries that were excluded for not fulfilling both criteria.

**Online supplementary methods: <u>Detailed search string</u>**

### A. MEDLINE

#### 1. Pathogen

exp Rotavirus Infections/ or exp ROTAVIRUS/

or

(rotavirus*2 or "rota virus*2").ti,ab,kw.

#### 2. Incidence

exp Disease Outbreaks/ or exp Communicable Diseases/ep or exp Communicable Diseases, Emerging/

or

(outbreak*1 or epidemic*1 or surveillance or gastroenteritis or diarrh?ea or stool or feces or morbidit*3 or "burden of disease" or "disease burden" or "primary care" or outpatient* or inpatient* or hospital* or nosocomial* or mortality or death* or incidence* or prevalence* or population-based).ti,ab,kw.

#### 3. Geography

exp European Union/ or exp EUROPE/ or

exp AUSTRALIA/ or exp AUSTRIA/ or exp BELGIUM/ or exp CANADA/ or exp Czech Republic/ or exp DENMARK/ or exp FINLAND/ or exp FRANCE/ or exp GERMANY/ or exp GREECE/ or exp ICELAND/ or exp IRELAND/ or exp ISRAEL/ or exp ITALY/ or exp JAPAN/ or exp LUXEMBOURG/ or exp NETHERLANDS/ or exp New Zealand/ or exp NORWAY/ or exp PORTUGAL/ or exp SLOVENIA/ or exp SPAIN/ or exp SWEDEN/ or exp SWITZERLAND/ or exp United Kingdom/ or exp United States/

or

(Europe* or "European Union" or "European Community" or EU or

Australia* or Sydney or Melbourne or Brisbane or Perth or Adelaide or "Gold Coast" or Newcastle or Canberra or

Austria or *austro* or Wien or Vienn* or Graz or Linz or Salzburg or

Belgium or Belgian* or Antwerp* or Ghent or Brussels or

Canad* or Toronto or Montreal or Calgary or Ottawa or Edmonton or Winnipeg or Vancouver or Quebec or

Czech* or Prague* or Praha or

Denmark or Danish or Copenhagen or Aarhus or Greenland or

Finland or Finnish or Finn*1 or Helsinki or

France or French or Paris or Marseille or Lyon or Toulouse or Nice or Nantes or Strasbourg or Montpellier or Lille or Bordeaux or

German* or Berlin or Hamburg or Munich or M#?nchen or Cologne or K#?ln or Frankfurt or Stuttgart or D#?sseldorf or Dortmund or

Greece or Greek* or Athen* or Thessaloniki or

Iceland* or Reykjavik or

Ireland or Irish or Eire or Dublin* or

Israel* or Jerusalem or "Tel Aviv" or Haifa or

Italy or Italian* or Rome or Roma* or Milan* or Naples or Napoli or Turin or Torino or Sicily or Genoa or Bologna or Firenze or Florence or

Japan* or Tokyo or Yokohama or Osaka or Nagoya or Sapporo or Fukuoka or Kobe or Kyoto or

Luxemb?urg* or

Netherland* or Holland or Dutch or Amsterdam or Rotterdam or Hague or Utrecht or Eindhoven or

"New Zealand*" or Auckland or Wellington or Christchurch or Hamilton or Dunedin or

Norway* or Norwegian* or Oslo or Bergen or Trondheim or Svalbard or

Portuguese* or Portugal or Lisbon or Porto or

Slovenia* or Ljubljana or

Spain or Spanish or Iberia* or Iberica* or Madrid or Barcelona or Valencia or Sevill* or Bilbao or M#laga or Zaragoza or Mallorca or

Sweden or Swedish or swede* or Stockholm or Gothenburg or Malm# or Uppsala or Norland or Svealand or Gotaland or

Schweiz or Suisse or Svizzera or Switzerland or Z#?rich or Zurich or Zurigo or Bern*1 or Luzern or Lucern*1 or Uri or Schwy?z or Svitto or Obwald*2 or Obvaldo or Nidwald*2 or Nidvaldo or Glarus or Glarona or Zug or Zoug or Zugo or Freiburg or Fribourg or Friburgo or Solothurn or Soleure or Soletta or Basel or B#le or Basilea or Basle or Schaffhausen or Schaffhouse or Sciaffusa or Appenzell* or "St. Gallen" or "Sankt Gallen" or Saint-Gall* or San Gallo or Graub#?nden or Grisons or Grigioni or Aargau or Argovie or Argovia or Thurgau or Thurgovie or Turgovia or Tessin or Tessin or Ticino or Waadt or Vaud or Wallis or Valais or Vallese or Neuenburg or Neuch#tel or Genf or Gen#ve or Ginevra or Geneva or Jura or Jura or Giura or

"United Kingdom" or UK or Britain or GB or British or England or English or Scotland or Scottish or Scots or Wales or Welsh or "Northern Ireland" or London or Manchester or Birmingham or Leeds or Liverpool or Glasgow or Edinburgh or Aberdeen or Cardiff or Belfast or

US or USA or "United States" or "United States of America" or Alabama or Georgia or Kentucky or Maryland or "New York" or "North Carolina" or Ohio or Pennsylvania or "South Carolina" or Tennessee or Virginia or "West Virginia" or Illinois or Indiana or Michigan or Minnesota or Ohio or Wisconsin or Delaware or "District of Columbia" or "New Jersey" or Iowa or Kansas or Kentucky or Missouri or Nebraska or "North Dakota" or Oklahoma or "South Dakota" or Wisconsin or "New England" or Connecticut or Maine or Massachusetts or "New Hampshire" or "Rhode Island" or Vermont or Idaho or Montana or Oregon or Washington or Wyoming or Alaska or California or Arkansas or Florida or Georgia or Louisiana or Mississippi or Arizona or Colorado or Nevada or "New Mexico" or Texas or Utah or "Los Angeles" or Chicago or Houston or Phoenix or Philadelphia or "San Antonio" or "San Diego" or Dallas or "San Francisco" or Seattle or Denver or Boston or Baltimore or "New Orleans" or global or developed or industriali#ed).ti,ab,kw,ot.

**4. Age category**

exp Child/ or exp Infant/ or exp Pediatrics/

or

(child* or infan* or p?ediatri*).ti,ab,kw,jn.

**5. Combine searches**

1 and 2 and 3 and 4

**6. Animal studies**

exp Animals/ not Humans/

**7. Combine searches**

5 not 6

### B. EMBASE

**1. Pathogen**

exp Rotavirus/

or

(rotavirus*2 or "rota virus*2").ti,ab,kw.

**2. Incidence**

exp epidemic/ or exp communicable disease/

or

(outbreak*1 or epidemic*1 or surveillance or gastroenteritis or diarrh?ea or stool or feces or morbidit*3 or "burden of disease" or "disease burden" or "primary care" or outpatient* or inpatient* or hospital* or nosocomial* or mortality or death* or incidence* or prevalence* or population-based).ti,ab,kw.

**3. Geography**

exp European Union/ or exp EUROPE/ or

exp Australia and New Zealand/ or exp Austria/ or exp Belgium/ or exp Canada/ or exp Czech Republic/ or exp Denmark/ or exp Finland/ or exp France/ or exp Germany/ or exp Greece/ or exp Iceland/ or exp Ireland/ or exp Israel/ or exp Italy/ or exp Japan/ or exp Luxembourg/ or exp Netherlands/ or exp Norway/ or exp Portugal/ or exp Slovenia/ or exp Spain/ or exp Sweden/ or exp Switzerland/ or exp United Kingdom/ or exp United States/

or

(Europe* or "European Union" or "European Community" or EU or

Australia* or Sydney or Melbourne or Brisbane or Perth or Adelaide or "Gold Coast" or Newcastle or Canberra or

Austria or *austro* or Wien or Vienn* or Graz or Linz or Salzburg or

Belgium or Belgian* or Antwerp* or Ghent or Brussels or

Canad* or Toronto or Montreal or Calgary or Ottawa or Edmonton or Winnipeg or Vancouver or Quebec or

Czech* or Prague* or Praha or

Denmark or Danish or Copenhagen or Aarhus or Greenland or

Finland or Finnish or Finn*1 or Helsinki or

France or French or Paris or Marseille or Lyon or Toulouse or Nice or Nantes or Strasbourg or Montpellier or Lille or Bordeaux or

German* or Berlin or Hamburg or Munich or M#?nchen or Cologne or K#?ln or Frankfurt or Stuttgart or D#?sseldorf or Dortmund or

Greece or Greek* or Athen* or Thessaloniki or

Iceland* or Reykjavik or

Ireland or Irish or Eire or Dublin* or

Israel* or Jerusalem or "Tel Aviv" or Haifa or

Italy or Italian* or Rome or Roma* or Milan* or Naples or Napoli or Turin or Torino or Sicily or Genoa or Bologna or Firenze or Florence or

Japan* or Tokyo or Yokohama or Osaka or Nagoya or Sapporo or Fukuoka or Kobe or Kyoto or

Luxemb?urg* or

Netherland* or Holland or Dutch or Amsterdam or Rotterdam or Hague or Utrecht or Eindhoven or

"New Zealand*" or Auckland or Wellington or Christchurch or Hamilton or Dunedin or

Norway* or Norwegian* or Oslo or Bergen or Trondheim or Svalbard or

Portuguese* or Portugal or Lisbon or Porto or

Slovenia* or Ljubljana or

Spain or Spanish or Iberia* or Iberica* or Madrid or Barcelona or Valencia or Sevill* or Bilbao or M#laga or Zaragoza or Mallorca or

Sweden or Swedish or swede* or Stockholm or Gothenburg or Malm# or Uppsala or Norland or Svealand or Gotaland or

Schweiz or Suisse or Svizzera or Switzerland or Z#?rich or Zurich or Zurigo or Bern*1 or Luzern or Lucern*1 or Uri or Schwy?z or Svitto or Obwald*2 or Obvaldo or Nidwald*2 or Nidvaldo or Glarus or Glarona or Zug or Zoug or Zugo or Freiburg or Fribourg or Friburgo or Solothurn or Soleure or Soletta or Basel or B#le or Basilea or Basle or Schaffhausen or Schaffhouse or Sciaffusa or Appenzell* or "St. Gallen" or "Sankt Gallen" or Saint-Gall* or San Gallo or Graub#?nden or Grisons or Grigioni or Aargau or Argovie or Argovia or Thurgau or Thurgovie or Turgovia or Tessin or Tessin or Ticino or Waadt or Vaud or Wallis or Valais or Vallese or Neuenburg or Neuch#tel or Genf or Gen#ve or Ginevra or Geneva or Jura or Jura or Giura or

"United Kingdom" or UK or Britain or GB or British or England or English or Scotland or Scottish or Scots or Wales or Welsh or "Northern Ireland" or London or Manchester or Birmingham or Leeds or Liverpool or Glasgow or Edinburgh or Aberdeen or Cardiff or Belfast or

US or USA or "United States" or "United States of America" or Alabama or Georgia or Kentucky or Maryland or "New York" or "North Carolina" or Ohio or Pennsylvania or "South Carolina" or Tennessee or Virginia or "West Virginia" or Illinois or Indiana or Michigan or Minnesota or Ohio or Wisconsin or Delaware or "District of Columbia" or "New Jersey" or Iowa or Kansas or Kentucky or Missouri or Nebraska or "North Dakota" or Oklahoma or "South Dakota" or Wisconsin or "New England" or Connecticut or Maine or Massachusetts or "New Hampshire" or "Rhode Island" or Vermont or Idaho or Montana or Oregon or Washington or Wyoming or Alaska or California or Arkansas or Florida or Georgia or Louisiana or Mississippi or Arizona or Colorado or Nevada or "New Mexico" or Texas or Utah or "Los Angeles" or Chicago or Houston or Phoenix or Philadelphia or "San Antonio" or "San Diego" or Dallas or "San Francisco" or Seattle or Denver or Boston or Baltimore or "New Orleans" or

global or developed or industriali#ed).ti,ab,kw,ot.

**4. Age category**

exp Child/ or exp Infant/ or exp Pediatrics/

or

(child* or infan* or p?ediatri*).ti,ab,kw,jn.

**5. Combine searches**

1 and 2 and 3 and 4

**6. Animal studies**

exp animal/ not human/

**7. Combine searches**

5 not 6

**Online supplementary methods: <u>Detailed items extracted</u>**

**Extraction and publication information:** Date of extraction, name of reviewer, title of the article, first author, email of corresponding author, year of publication, journal, volume, issue and pages, language of the article.

**Study characteristics:** study design, unique identifier (ID), clinical or population-based, prospective or retrospective data retrieval (health records or data linkage), country of study, level of urbanisation, single centre or multi-centre, regional, national or international study, length of study or follow up period, months and year(s) of recorded events, ethical approval.

**Characteristics of participants:** sample selection, number of participants (absolute and person-years), reference population (number), age range, gender, method of recruitment (e.g. hospital, primary care, etc.), response rate.

**Details of outcomes reported:** Specific outcome measure used (primary care visit, emergency care visit, hospitalisation, nosocomial infection or death), definition of the outcome measure, identification of causal pathogen for the gastroenteritis (specific laboratory procedure), results for each outcome measure (absolute number, proportion, rate) with 95% CI.

**Supplementary Figure 1.** Proportion of hospitalisations for RVGE among all hospitalisations for acute gastroenteritis of children aged 0-5 years (forest plot with country estimates)

| Reference | Study period | Age (months) | No. of RV+ | Total AGE | 95%-CI | Weight |
|---|---|---|---|---|---|---|
| **Belgium** | | | | | | |
| Raes 2007 | 06/02-05/06 | 0-59 | 423 | 1074 | 0.39 [0.36; 0.42] | 2.2% |
| Raes 2011 | 06/04-05/06 | 0-59 | 881 | 3161 | 0.28 [0.26; 0.29] | 2.2% |
| Random effects model | | | | 4235 | 0.33 [0.23; 0.45] | 4.3% |
| Heterogeneity: $I^2$ = 98%, $\tau^2$ = 0.0073, $p$ < 0.01 | | | | | | |
| **Czech Republic** | | | | | | |
| Arientova 2012 | 09/06-12/08 | 0-59 | 478 | 832 | 0.57 [0.54; 0.61] | 2.2% |
| Random effects model | | | | 832 | 0.57 [0.54; 0.61] | 2.2% |
| Heterogeneity: not applicable | | | | | | |
| **Denmark** | | | | | | |
| Fischer 2001 | 01/95-12/99 | 0-59 | 434 | 2146 | 0.20 [0.19; 0.22] | 2.2% |
| Fischer 2011 | 02/09-03/10 | 0-59 | 1074 | 2610 | 0.41 [0.39; 0.43] | 2.2% |
| Random effects model | | | | 4756 | 0.30 [0.12; 0.52] | 4.3% |
| Heterogeneity: $I^2$ = 100%, $\tau^2$ = 0.0263, $p$ < 0.01 | | | | | | |
| **Europe** | | | | | | |
| Giaquinto 2007 | 10/04-09/05 | 0-59 | 337 | 555 | 0.61 [0.57; 0.65] | 2.1% |
| Random effects model | | | | 555 | 0.61 [0.57; 0.65] | 2.1% |
| Heterogeneity: not applicable | | | | | | |
| **France** | | | | | | |
| Forster 2009 | 02/05-08/06 | 0-59 | 186 | 289 | 0.64 [0.59; 0.70] | 2.1% |
| Parez 2011 | 05/05-05/06 | 0-59 | 186 | 289 | 0.64 [0.59; 0.70] | 2.1% |
| Random effects model | | | | 578 | 0.64 [0.60; 0.68] | 4.2% |
| Heterogeneity: $I^2$ = 0%, $\tau^2$ = 0, $p$ = 1.00 | | | | | | |
| **Germany** | | | | | | |
| Poppe 2002 | 06/97-05/98 | 0-48 | 621 | 1451 | 0.43 [0.40; 0.45] | 2.2% |
| Forster 2009 | 02/05-08/06 | 0-59 | 380 | 621 | 0.61 [0.57; 0.65] | 2.1% |
| Kaiser 2011 | 10/02-05/08 | 0-59 | 2118 | 4867 | 0.44 [0.42; 0.45] | 2.2% |
| Koch 2011 | 01/01-12/08 | 0-59 | 17636 | 25154 | 0.70 [0.70; 0.71] | 2.2% |
| Pietsch 2018 | 10/02-09/17 | 0-59 | 1024 | 3226 | 0.32 [0.30; 0.33] | 2.2% |
| Random effects model | | | | 35319 | 0.50 [0.32; 0.68] | 10.8% |
| Heterogeneity: $I^2$ = 100%, $\tau^2$ = 0.0436, $p$ = 0 | | | | | | |
| **Greece** | | | | | | |
| Levidioutou 2009 | 01/00-12/06 | 0-59 | 904 | 4604 | 0.20 [0.18; 0.21] | 2.2% |
| Nika 2010 | 01/08-12/09 | 0-59 | 216 | 1352 | 0.16 [0.14; 0.18] | 2.2% |
| Konstantopoulos 2013 | 07/08-03/10 | 0-59 | 109 | 458 | 0.24 [0.20; 0.28] | 2.1% |
| Random effects model | | | | 6414 | 0.19 [0.16; 0.23] | 6.5% |
| Heterogeneity: $I^2$ = 88%, $\tau^2$ = 0.0013, $p$ < 0.01 | | | | | | |
| **Israel** | | | | | | |
| Givon-Lavi 2008 | 04/06-03/07 | 0-59 | 176 | 568 | 0.31 [0.27; 0.35] | 2.1% |
| Muhsen 2015 | 11/07-07/14 | 0-59 | 1137 | 3514 | 0.32 [0.31; 0.34] | 2.2% |
| Random effects model | | | | 4082 | 0.32 [0.31; 0.34] | 4.3% |
| Heterogeneity: $I^2$ = 0%, $\tau^2$ = 0, $p$ = 0.52 | | | | | | |

Proportion RV+ (%)

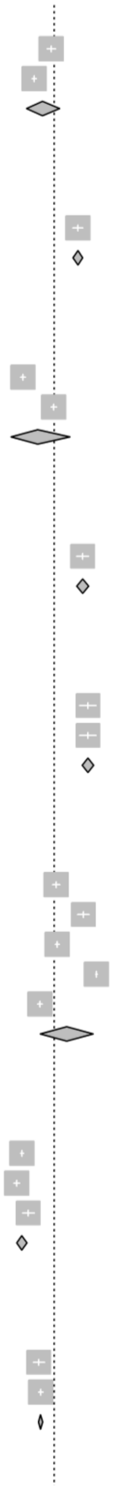

*Supplementary Figure 1 (continued)*

| Reference | Study period | Age (months) | No. of RV+ | Total AGE | 95%-CI | Weight |
|---|---|---|---|---|---|---|
| **Italy** | | | | | | |
| Albano 2006 | NA-NA | 1-48 | 381 | 911 | 0.42 [0.39; 0.45] | 2.2% |
| De Grazia 2006 | 01/01-12/05 | 0-59 | 411 | 1030 | 0.40 [0.37; 0.43] | 2.2% |
| Giaquinto 2008 | 10/04-09/05 | 1-59 | 55 | 83 | 0.66 [0.55; 0.76] | 2.0% |
| Panatto 2009 | 01/06-12/06 | 0-59 | 216 | 776 | 0.28 [0.25; 0.31] | 2.2% |
| Forster 2009 | 02/05-08/06 | 0-59 | 66 | 199 | 0.33 [0.27; 0.40] | 2.1% |
| Zuccotti 2010 | 01/08-12/08 | 0-59 | 182 | 521 | 0.35 [0.31; 0.39] | 2.1% |
| De Grazia 2014 | 01/85-08/12 | 0-59 | 2138 | 6522 | 0.33 [0.32; 0.34] | 2.2% |
| Dona 2016 | 05/10-04/11 | 0-59 | 3 | 12 | 0.25 [0.05; 0.57] | 1.4% |
| Random effects model | | | | 10054 | 0.37 [0.33; 0.42] | 16.3% |
| Heterogeneity: $I^2 = 92\%$, $\tau^2 = 0.0039$, $p < 0.01$ | | | | | | |
| **Spain** | | | | | | |
| Sanchez-Fauquier 2003 | 10/98-09/02 | 0-59 | 1155 | 3760 | 0.31 [0.29; 0.32] | 2.2% |
| Sanchez-Fauquier 2006 | 01/05-12/05 | 0-59 | 362 | 656 | 0.55 [0.51; 0.59] | 2.1% |
| Forster 2009 | 02/05-08/06 | 0-59 | 86 | 164 | 0.52 [0.45; 0.60] | 2.1% |
| Cilla 2010 | 07/96-06/08 | 1-59 | 419 | 1070 | 0.39 [0.36; 0.42] | 2.2% |
| Sanchez-Fauquier 2011 | 03/06-04/08 | 0-59 | 821 | 2048 | 0.40 [0.38; 0.42] | 2.2% |
| Rivero 2011 | 01/05-12/08 | 0-59 | 323 | 663 | 0.49 [0.45; 0.53] | 2.1% |
| Castilla 2011 | 01/08-06/11 | 0-59 | 262 | 780 | 0.34 [0.30; 0.37] | 2.2% |
| Gonzalez Galan 2011 | 04/06-04/07 | 0-59 | 80 | 399 | 0.20 [0.16; 0.24] | 2.1% |
| Random effects model | | | | 9540 | 0.40 [0.33; 0.46] | 17.2% |
| Heterogeneity: $I^2 = 97\%$, $\tau^2 = 0.0093$, $p < 0.01$ | | | | | | |
| **Sweden** | | | | | | |
| Rinder 2014 | 10/07-10/08 | 0-59 | 642 | 1549 | 0.41 [0.39; 0.44] | 2.2% |
| Random effects model | | | | 1549 | 0.41 [0.39; 0.44] | 2.2% |
| Heterogeneity: not applicable | | | | | | |
| **United Kingdom** | | | | | | |
| Forster 2009 | 02/05-08/06 | 0-59 | 27 | 52 | 0.52 [0.38; 0.66] | 1.9% |
| Random effects model | | | | 52 | 0.52 [0.38; 0.65] | 1.9% |
| Heterogeneity: not applicable | | | | | | |
| **Australia** | | | | | | |
| Schnagl 2002 | 01/95-12/98 | 1-59 | 267 | 774 | 0.34 [0.31; 0.38] | 2.2% |
| Galati 2006 | 07/98-06/03 | 0-59 | 2344 | 4688 | 0.50 [0.49; 0.51] | 2.2% |
| Campbell 2009 | 07/01-06/06 | 0-59 | 310 | 881 | 0.35 [0.32; 0.38] | 2.2% |
| Jayasinghe 2012 | 01/00-12/07 | 0-59 | 689 | 1453 | 0.47 [0.45; 0.50] | 2.2% |
| Random effects model | | | | 7796 | 0.42 [0.34; 0.50] | 8.6% |
| Heterogeneity: $I^2 = 97\%$, $\tau^2 = 0.0063$, $p < 0.01$ | | | | | | |
| **Japan** | | | | | | |
| Kamiya 2009 | 01/03-12/07 | 0-59 | 453 | 961 | 0.47 [0.44; 0.50] | 2.2% |
| Tajiri 2013 | 01/08-12/09 | 0-59 | 625 | 1072 | 0.58 [0.55; 0.61] | 2.2% |
| Kinoshita 2014 | 09/01-08/11 | 0-59 | 387 | 834 | 0.46 [0.43; 0.50] | 2.2% |
| Random effects model | | | | 2867 | 0.51 [0.43; 0.58] | 6.5% |
| Heterogeneity: $I^2 = 94\%$, $\tau^2 = 0.0044$, $p < 0.01$ | | | | | | |
| **New Zealand** | | | | | | |
| Kelly 2017 | 01/10-12/14 | 0-59 | 401 | 906 | 0.44 [0.41; 0.48] | 2.2% |
| Random effects model | | | | 906 | 0.44 [0.41; 0.48] | 2.2% |
| Heterogeneity: not applicable | | | | | | |
| **United States** | | | | | | |
| Lee 2005 | 11/97-12/99 | 0-59 | 548 | 2067 | 0.27 [0.25; 0.28] | 2.2% |
| Patel 2007 | 07/00-06/06 | 0-59 | 1109 | 1802 | 0.62 [0.59; 0.64] | 2.2% |
| Mast 2009 | 02/05-07/06 | 0-59 | 201 | 411 | 0.49 [0.44; 0.54] | 2.1% |
| Random effects model | | | | 4280 | 0.45 [0.22; 0.70] | 6.5% |
| Heterogeneity: $I^2 = 100\%$, $\tau^2 = 0.0509$, $p < 0.01$ | | | | | | |
| **Random effects model** | | | | **93815** | **0.41 [0.36; 0.47]** | **100.0%** |
| **Prediction interval** | | | | | **[0.09; 0.79]** | |
| Heterogeneity: $I^2 = 100\%$, $\tau^2 = 0.0363$, $p = 0$ | | | | | | |
| Residual heterogeneity: $I^2 = 99\%$, $p = 0$ | | | | | | |

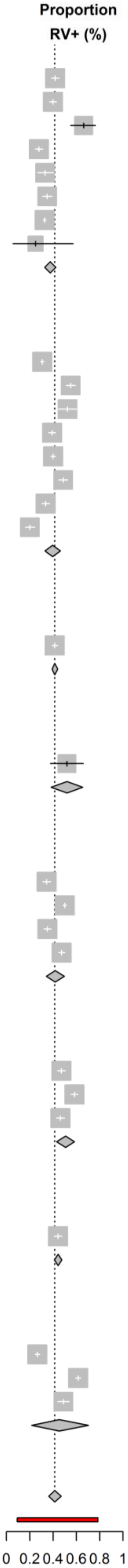


RVGE: Rotavirus gastroenteritis; RV+: stools samples that tested positive for rotavirus; AGE: acute gastroenteritis; CI: Confidence Interval

**Supplementary Figure 2.** Incidence rate of RVGE nosocomial infections among children aged 0-5 years hospitalised for reasons other than acute gastroenteritis per 100 000 person-years (forest plot)

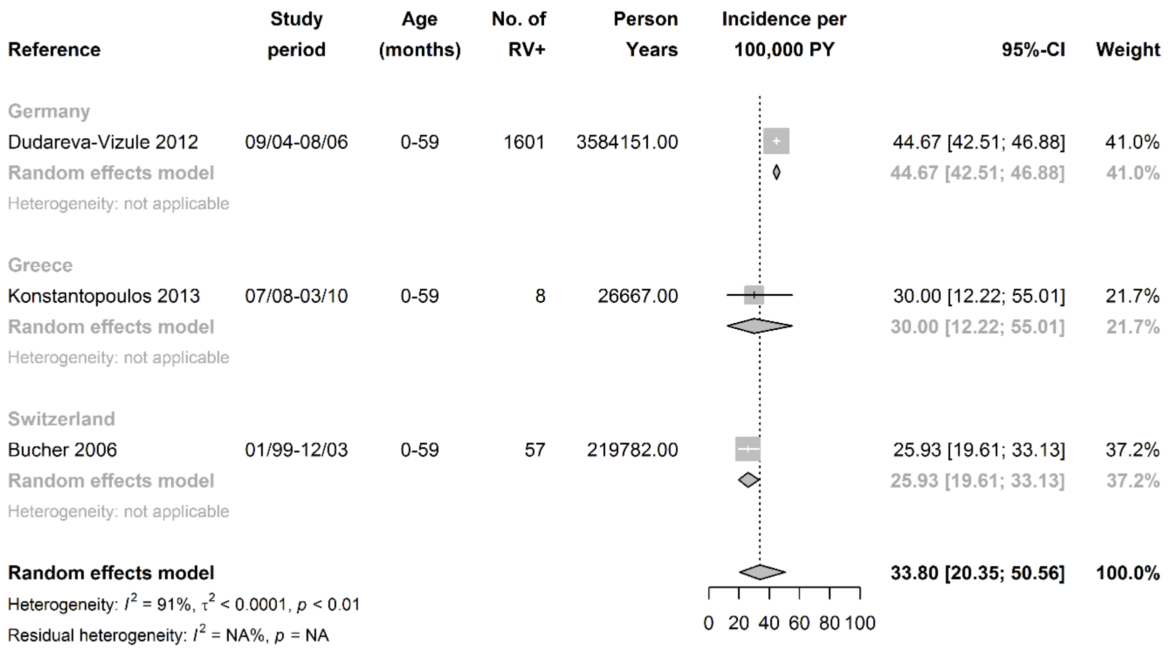


RVGE: Rotavirus gastroenteritis; RV+: stools samples that tested positive for rotavirus; PY: person-years; CI: Confidence Interval

**Supplementary Figure 3.** Incidence rate of RVGE nosocomial infections among children aged 0-5 years hospitalised for reasons other than acute gastroenteritis per 1000 hospital-days (forest plot)

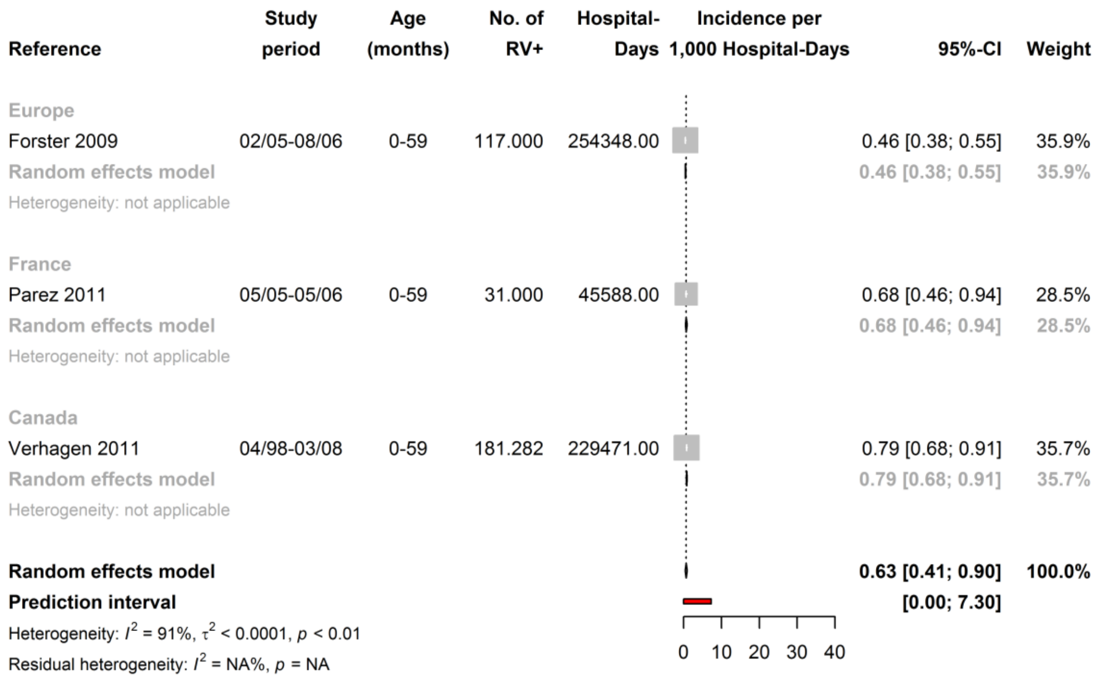


RVGE: Rotavirus gastroenteritis; RV+: stools samples that tested positive for rotavirus; CI: Confidence Interval

**Supplementary Figure 4.** Proportion of acute gastroenteritis nosocomial infections caused by RV among children aged 0-5 years hospitalised for reasons other than acute gastroenteritis (forest plot)

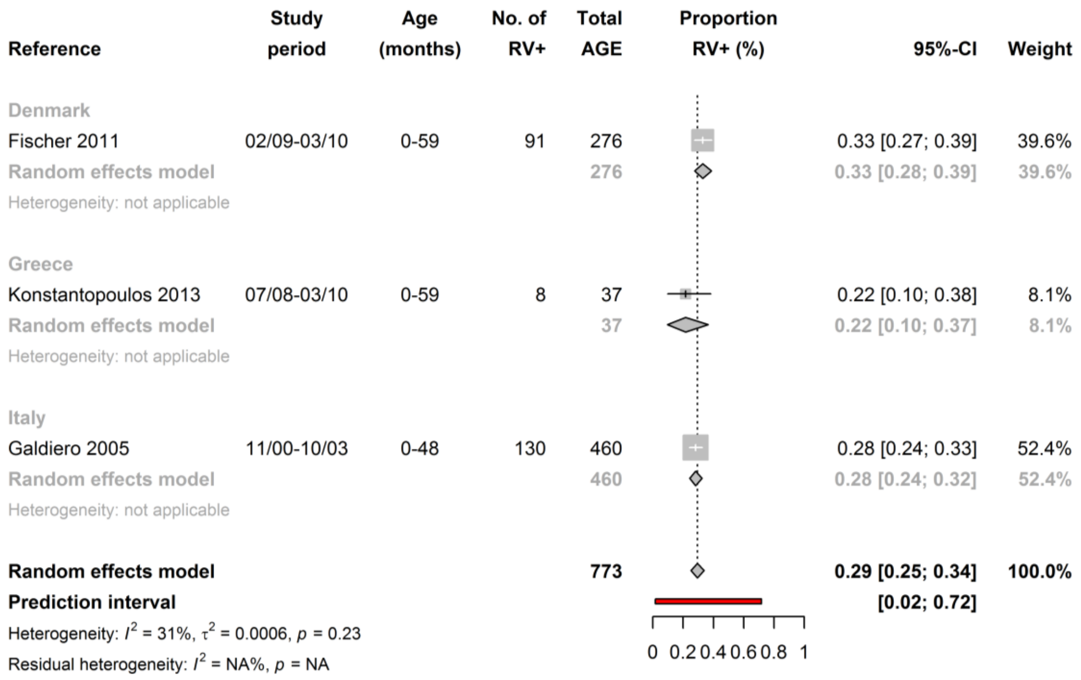


RVGE: Rotavirus gastroenteritis; RV+: stools samples that tested positive for rotavirus; AGE: acute gastroenteritis; CI: Confidence Interval

**Supplementary Figure 5**. Number of studies that fulfilled each item of Hoy et al. risk of bias tool (N= 74)

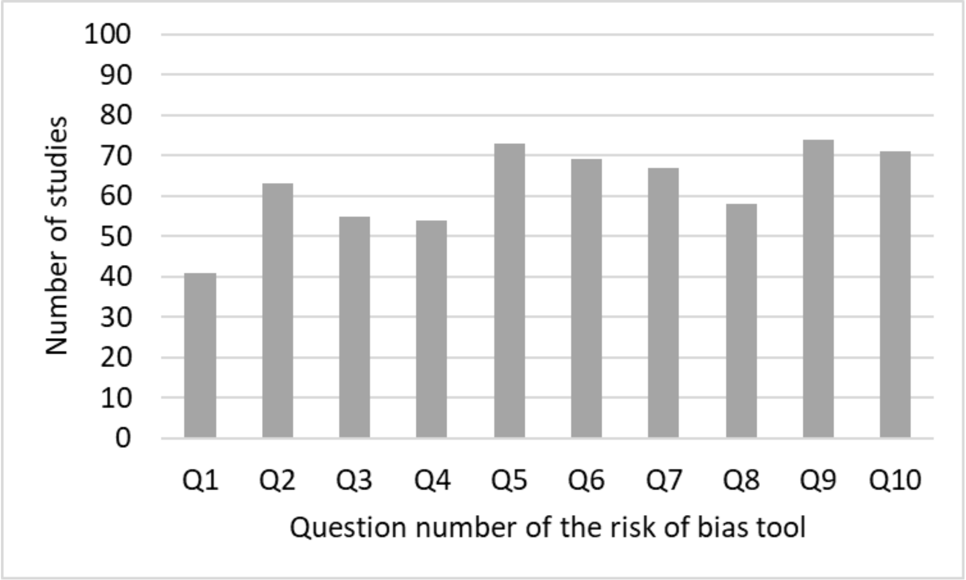

**Supplementary Figure 6**. Incidence rate of primary care visits for RVGE of children aged 0-2 years per 100 000 person-years (forest plot)

| Reference | Study period | Age (months) | No. of RV+ | Person Years | Incidence per 100,000 PY | 95%-CI | Weight |
|---|---|---|---|---|---|---|---|
| Finland | | | | | | | |
| Vesikari 2007 | 01/01-12/04 | 1-24 | 58 | 1568.00 | | 3698.98 [2804.21; 4716.25] | 55.6% |
| Random effects model | | | | | | 3698.98 [2804.21; 4716.25] | 55.6% |
| Heterogeneity: not applicable | | | | | | | |
| United States | | | | | | | |
| Vesikari 2007 | 01/01-12/04 | 1-24 | 40 | 748.00 | | 5347.59 [3808.57; 7143.43] | 44.4% |
| Random effects model | | | | | | 5347.59 [3808.57; 7143.43] | 44.4% |
| Heterogeneity: not applicable | | | | | | | |
| **Random effects model** | | | | | | **4390.57 [2916.48; 6161.10]** | **100.0%** |
| Heterogeneity: $I^2$ = 68%, $\tau^2$ = 0.0005, $p$ = 0.08 | | | | | | | |
| Residual heterogeneity: $I^2$ = NA%, $p$ = NA | | | | | | | |

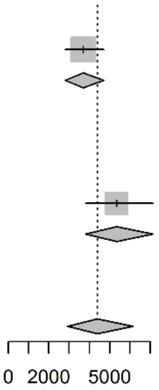


RVGE: Rotavirus gastroenteritis; RV+: stools samples that tested positive for rotavirus; PY: person-years; CI: Confidence Interval

**Supplementary Figure 7**. Proportion of primary care visits for RVGE among all visits for acute gastroenteritis of children aged 0-2 years (forest plot)

| Reference | Study period | Age (months) | No. of RV+ | Total AGE | Proportion RV+ (%) | 95%-CI | Weight |
|---|---|---|---|---|---|---|---|
| Italy | | | | | | | |
| Panatto 2009 | 01/06-12/06 | 0-23 | 34 | 113 | | 0.30 [0.22; 0.39] | 45.2% |
| Random effects model | | | | 113 | | 0.30 [0.22; 0.39] | 45.2% |
| Heterogeneity: not applicable | | | | | | | |
| United Kingdom | | | | | | | |
| Iturriza-Gomara 2008 | 02/06-09/07 | 0-24 | 92 | 487 | | 0.19 [0.16; 0.23] | 54.8% |
| Random effects model | | | | 487 | | 0.19 [0.16; 0.22] | 54.8% |
| Heterogeneity: not applicable | | | | | | | |
| **Random effects model** | | | | **600** | | **0.24 [0.14; 0.35]** | **100.0%** |
| Heterogeneity: $I^2$ = 84%, $\tau^2$ = 0.0074, $p$ = 0.01 | | | | | | | |
| Residual heterogeneity: $I^2$ = NA%, $p$ = NA | | | | | | | |

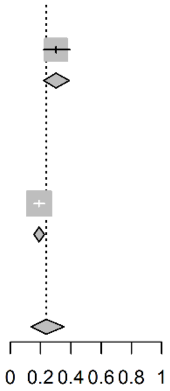


RVGE: Rotavirus gastroenteritis; RV+: stools samples that tested positive for rotavirus; AGE: acute gastroenteritis; CI: Confidence Interval

**Supplementary Figure 8.** Incidence rate of emergency department consultations for RVGE of children aged 0-2 years per 100 000 person-years (forest plot)

| Reference | Study period | Age (months) | No. of RV+ | Person Years | Incidence per 100,000 PY | 95%-CI | Weight |
|---|---|---|---|---|---|---|---|
| Europe | | | | | | | |
| Vesikari 2009 | 01/01-12/05 | 1-24 | 193 | 9415.00 | | 2049.92 [1770.54; 2349.70] | 50.1% |
| Random effects model | | | | | | 2049.92 [1770.54; 2349.70] | 50.1% |
| Heterogeneity: not applicable | | | | | | | |
| United States | | | | | | | |
| Vesikari 2007 | 01/01-12/04 | 1-24 | 42 | 7241.00 | | 580.03 [ 416.85; 769.74] | 49.9% |
| Random effects model | | | | | | 580.03 [ 416.85; 769.74] | 49.9% |
| Heterogeneity: not applicable | | | | | | | |
| **Random effects model** | | | | | | **1204.90 [ 193.13; 3072.49]** | **100.0%** |
| Heterogeneity: $I^2$ = 99%, $\tau^2$ = 0.0022, $p$ < 0.01 | | | | | | | |
| Residual heterogeneity: $I^2$ = NA%, $p$ = NA | | | | | | | |

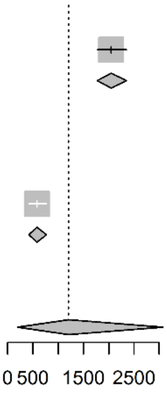


RVGE: Rotavirus gastroenteritis; RV+: stools samples that tested positive for rotavirus; PY: person-years; CI: Confidence Interval

**Supplementary Figure 9.** Proportion of emergency department consultations for RVGE among all consultations for acute gastroenteritis of children aged 0-2 years (forest plot)

| Reference | Study period | Age (months) | No. of RV+ | Total AGE | Proportion RV+ (%) | 95%-CI | Weight |
|---|---|---|---|---|---|---|---|
| France | | | | | | | |
| Parez 2011 | 05/05-05/06 | 0-23 | 96 | 189 | | 0.51 [0.43; 0.58] | 49.5% |
| Random effects model | | | | 189 | | 0.51 [0.44; 0.58] | 49.5% |
| Heterogeneity: not applicable | | | | | | | |
| Japan | | | | | | | |
| Thongprachum 2015 | 07/09-06/13 | 0-23 | 345 | 1715 | | 0.20 [0.18; 0.22] | 50.5% |
| Random effects model | | | | 1715 | | 0.20 [0.18; 0.22] | 50.5% |
| Heterogeneity: not applicable | | | | | | | |
| **Random effects model** | | | | **1904** | | **0.34 [0.09; 0.66]** | **100.0%** |
| Heterogeneity: $I^2$ = 99%, $\tau^2$ = 0.0531, $p$ < 0.01 | | | | | | | |
| Residual heterogeneity: $I^2$ = NA%, $p$ = NA | | | | | | | |

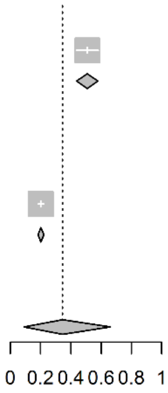


RVGE: Rotavirus gastroenteritis; RV+: stools samples that tested positive for rotavirus; PY: person-years; CI: Confidence Interval

**Supplementary Figure 10**. Incidence rate of hospitalisations for RVGE of children aged 0-2 years per 100 000 person-years (forest plot)

| Reference | Study period | Age (months) | No. of RV+ | Person Years | Incidence per 100,000 PY 95%-CI | Weight |
|---|---|---|---|---|---|---|
| **Austria** | | | | | | |
| Rendi-Wagner 2006 | 01/97-12/03 | 0-23 | 2498.0000 | 143398.00 | 1742.00 [1674.35; 1810.99] | 11.4% |
| **Random effects model** | | | | | **1742.00 [1674.35; 1810.99]** | **11.4%** |
| Heterogeneity: not applicable | | | | | | |
| **Europe** | | | | | | |
| Vesikari 2009 | 01/01-12/05 | 1-24 | 138.0000 | 9388.00 | 1469.96 [1234.50; 1725.89] | 10.3% |
| **Random effects model** | | | | | **1469.96 [1234.50; 1725.89]** | **10.3%** |
| Heterogeneity: not applicable | | | | | | |
| **Germany** | | | | | | |
| Koch 2011 | 01/01-12/08 | 0-23 | 13036.0000 | 1372959.00 | 949.48 [ 933.25; 965.85] | 11.5% |
| Wilking 2012 | 01/07-12/09 | 0-23 | 1511.0000 | 185649.00 | 813.90 [ 773.37; 855.46] | 11.4% |
| Dudareva-Vizule 2012 | 09/04-08/06 | 0-23 | 15049.0000 | 1393871.00 | 1079.66 [1062.47; 1096.97] | 11.5% |
| **Random effects model** | | | | | **946.33 [ 830.65; 1069.54]** | **34.3%** |
| Heterogeneity: $I^2 = 99\%$, $\tau^2 = < 0.0001$, $p < 0.01$ | | | | | | |
| **Ireland** | | | | | | |
| Lynch 2000 | 01/96-12/98 | 0-23 | 2504.0000 | 194856.00 | 1285.05 [1235.21; 1335.88] | 11.4% |
| **Random effects model** | | | | | **1285.05 [1235.21; 1335.88]** | **11.4%** |
| Heterogeneity: not applicable | | | | | | |
| **Switzerland** | | | | | | |
| Bucher 2006 | 01/99-12/03 | 0-23 | 214.0000 | 84005.00 | 254.75 [ 221.72; 290.06] | 11.3% |
| **Random effects model** | | | | | **254.75 [ 221.72; 290.06]** | **11.3%** |
| Heterogeneity: not applicable | | | | | | |
| **United States** | | | | | | |
| Vesikari 2007 | 01/01-12/04 | 1-24 | 16.0000 | 7273.00 | 219.99 [ 123.78; 342.63] | 10.0% |
| Staat 2010 | 07/96-06/08 | 0-23 | 205.3556 | 57901.00 | 354.67 [ 307.76; 404.89] | 11.3% |
| **Random effects model** | | | | | **297.30 [ 184.30; 436.77]** | **21.3%** |
| Heterogeneity: $I^2 = 73\%$, $\tau^2 = < 0.0001$, $p = 0.05$ | | | | | | |
| **Random effects model** | | | | | **826.95 [ 650.75; 1024.19]** | **100.0%** |
| Heterogeneity: $I^2 = 100\%$, $\tau^2 = 0.0002$, $p = 0$ | | | | | | |
| Residual heterogeneity: $I^2 = 98\%$, $p < 0.01$ | | | | | | |

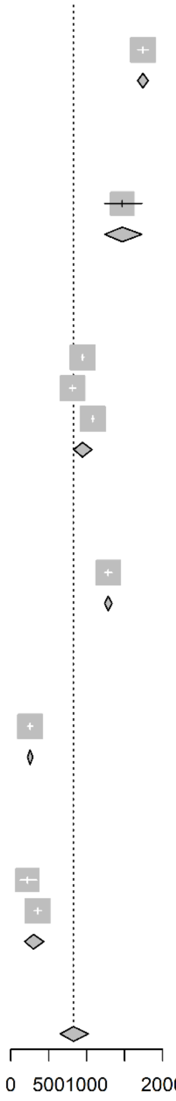


RVGE: Rotavirus gastroenteritis; RV+: stools samples that tested positive for rotavirus; PY: person-years; CI: Confidence Interval

**Supplementary Figure 11**. Proportion of hospitalisations for RVGE among all hospitalisations for acute gastroenteritis of children aged 0-2 years (forest plot)

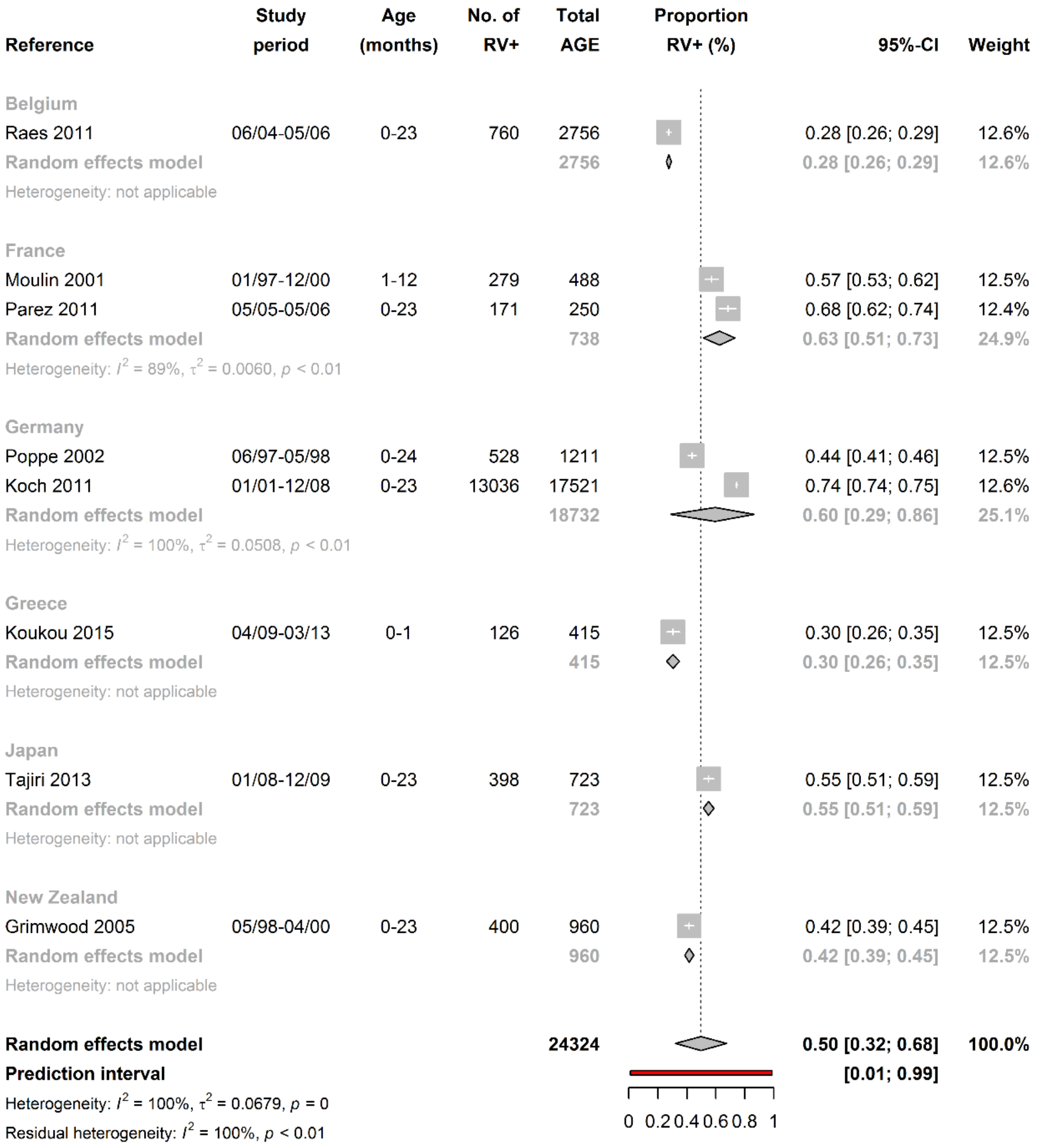


RVGE: Rotavirus gastroenteritis; RV+: stools samples that tested positive for rotavirus; AGE: acute gastroenteritis; CI: Confidence Interval

**Supplementary Figure 12.** Incidence proportion of RVGE nosocomial infections among children aged 0-2 years hospitalised for reasons other than acute gastroenteritis per 1000 hospitalisations (forest plot)

| Reference | Study period | Age (months) | No. of RV+ | Hospitalisations | Incidence per 1,000 Hospitalisations | 95%-CI | Weight |
|---|---|---|---|---|---|---|---|
| Spain | | | | | | | |
| Roman Riechmann 2003 | 06/98-05/99 | 1-24 | 34 | 666.00 | | 51.05 [35.21; 69.78] | 32.5% |
| Redondo-Gonzalez 2015 | 01/03-12/09 | 0-1 | 27 | 4018.00 | | 6.72 [ 4.40; 9.52] | 34.6% |
| Random effects model | | | | | | 23.30 [ 0.00; 87.20] | 67.0% |
| Heterogeneity: $I^2$ = 98%, $\tau^2$ = 0.0103, $p$ < 0.01 | | | | | | | |
| Switzerland | | | | | | | |
| Buettcher 2010 | 08/06-07/08 | 2-20 | 8 | 851.00 | | 9.40 [ 3.83; 17.24] | 33.0% |
| Random effects model | | | | | | 9.40 [ 3.83; 17.24] | 33.0% |
| Heterogeneity: not applicable | | | | | | | |
| **Random effects model** | | | | | | **17.85 [ 2.59; 45.98]** | **100.0%** |
| Heterogeneity: $I^2$ = 96%, $\tau^2$ = 0.0048, $p$ < 0.01 | | | | | | | |
| Residual heterogeneity: $I^2$ = 98%, $p$ < 0.01 | | | | | | | |

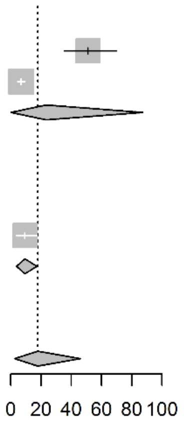


RVGE: Rotavirus gastroenteritis; RV+: stools samples that tested positive for rotavirus; CI: Confidence Interval

**Supplementary Figure 13.** Incidence rate of RVGE nosocomial infections among children aged 0-2 years hospitalised for reasons other than acute gastroenteritis per 1000 hospital-days (forest plot)

| Reference | Study period | Age (months) | No. of RV+ | Hospital-Days | Incidence per 1,000 Hospital-Days | 95%-CI | Weight |
|---|---|---|---|---|---|---|---|
| France | | | | | | | |
| Marc 2006 | 11/97-02/01 | 0-24 | 64 | 2826.00 | | 22.65 [17.42; 28.56] | 24.7% |
| Random effects model | | | | | | 22.65 [17.42; 28.56] | 24.7% |
| Heterogeneity: not applicable | | | | | | | |
| Spain | | | | | | | |
| Roman Riechmann 2003 | 06/98-05/99 | 1-24 | 34 | 3429.00 | | 9.92 [ 6.84; 13.55] | 24.8% |
| Redondo-Gonzalez 2015 | 01/03-12/09 | 0-1 | 27 | 13333.00 | | 2.03 [ 1.33; 2.87] | 25.5% |
| Random effects model | | | | | | 5.15 [ 0.26; 15.82] | 50.3% |
| Heterogeneity: $I^2$ = 97%, $\tau^2$ = 0.0015, $p$ < 0.01 | | | | | | | |
| Switzerland | | | | | | | |
| Buettcher 2010 | 08/06-07/08 | 2-20 | 8 | 4255.00 | | 1.88 [ 0.77; 3.45] | 25.0% |
| Random effects model | | | | | | 1.88 [ 0.77; 3.45] | 25.0% |
| Heterogeneity: not applicable | | | | | | | |
| **Random effects model** | | | | | | **7.09 [ 1.42; 16.95]** | **100.0%** |
| Heterogeneity: $I^2$ = 98%, $\tau^2$ = 0.0021, $p$ < 0.01 | | | | | | | |
| Residual heterogeneity: $I^2$ = 97%, $p$ < 0.01 | | | | | | | |

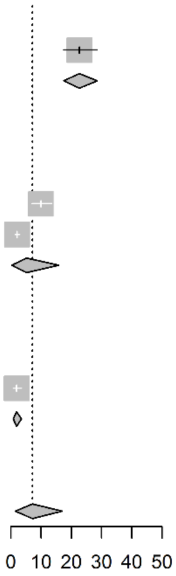


RVGE: Rotavirus gastroenteritis; RV+: stools samples that tested positive for rotavirus; CI: Confidence Interval

**Supplementary Figure 14**. Proportion of acute gastroenteritis nosocomial infections caused by RV among children aged 0-2 years hospitalised for reasons other than acute gastroenteritis (forest plot)

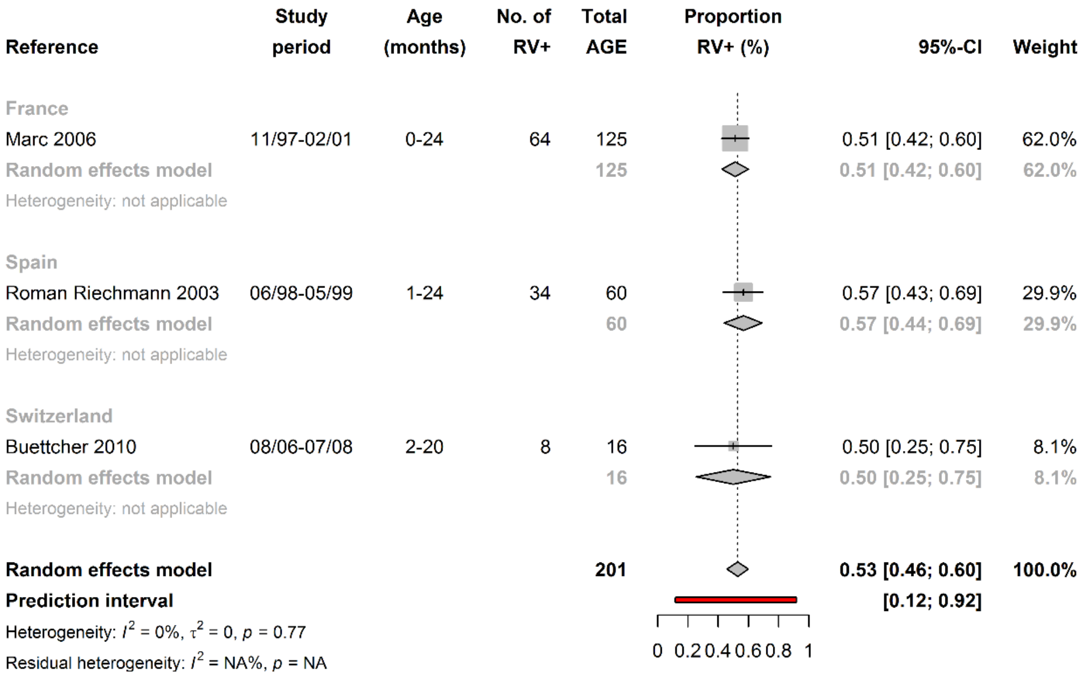


RVGE: Rotavirus gastroenteritis; RV+: stools samples that tested positive for rotavirus; AGE: acute gastroenteritis; CI: Confidence Interval

**Supplementary Figure 15.** Proportion of primary care visits for RVGE among all visits for acute gastroenteritis of children aged 0-5 years, including only studies with good external validity (forest plot)

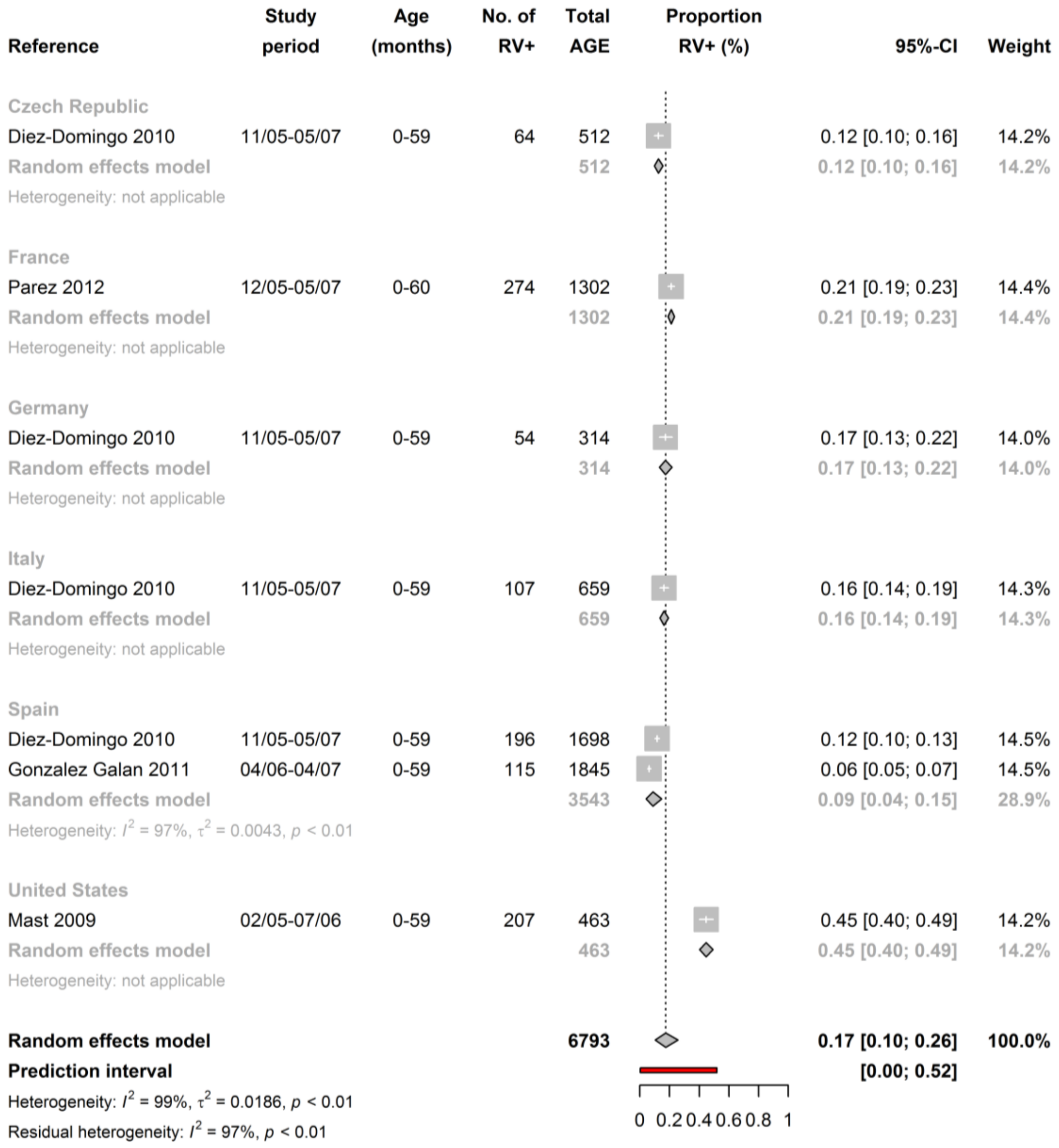


RVGE: Rotavirus gastroenteritis; RV+: stools samples that tested positive for rotavirus; AGE: acute gastroenteritis; CI: Confidence Interval

**Supplementary Figure 16**. Proportion of emergency department consultations for RVGE among all consultations for acute gastroenteritis of children aged 0-5 years, including only studies with good external validity (forest plot)

| Reference | Study period | Age (months) | No. of RV+ | Total AGE | 95%-CI | Weight |
|---|---|---|---|---|---|---|
| France | | | | | | |
| Forster 2009 | 02/05-08/06 | 0-59 | 114 | 232 | 0.49 [0.43; 0.56] | 9.1% |
| Random effects model | | | | 232 | 0.49 [0.43; 0.56] | 9.1% |
| Heterogeneity: not applicable | | | | | | |
| Germany | | | | | | |
| Forster 2009 | 02/05-08/06 | 0-59 | 114 | 278 | 0.41 [0.35; 0.47] | 9.1% |
| Random effects model | | | | 278 | 0.41 [0.35; 0.47] | 9.1% |
| Heterogeneity: not applicable | | | | | | |
| Greece | | | | | | |
| Konstantopoulos 2013 | 07/08-03/10 | 0-59 | 11 | 103 | 0.11 [0.05; 0.18] | 8.7% |
| Random effects model | | | | 103 | 0.11 [0.05; 0.17] | 8.7% |
| Heterogeneity: not applicable | | | | | | |
| Italy | | | | | | |
| Forster 2009 | 02/05-08/06 | 0-59 | 126 | 382 | 0.33 [0.28; 0.38] | 9.2% |
| Random effects model | | | | 382 | 0.33 [0.28; 0.38] | 9.2% |
| Heterogeneity: not applicable | | | | | | |
| Spain | | | | | | |
| Dalton 2002 | 10/96-09/97 | 0-48 | 205 | 822 | 0.25 [0.22; 0.28] | 9.3% |
| Forster 2009 | 02/05-08/06 | 0-59 | 160 | 661 | 0.24 [0.21; 0.28] | 9.3% |
| Random effects model | | | | 1483 | 0.25 [0.22; 0.27] | 18.6% |
| Heterogeneity: $I^2 = 0\%$, $\tau^2 = 0$, $p = 0.75$ | | | | | | |
| United Kingdom | | | | | | |
| Forster 2009 | 02/05-08/06 | 0-59 | 12 | 50 | 0.24 [0.13; 0.38] | 8.1% |
| Random effects model | | | | 50 | 0.24 [0.13; 0.37] | 8.1% |
| Heterogeneity: not applicable | | | | | | |
| Canada | | | | | | |
| Comeau 2016 | 07/06-06/13 | 0-59 | 432 | 2944 | 0.15 [0.13; 0.16] | 9.4% |
| Freedman 2018 | 11/13-04/17 | 3-48 | 209 | 860 | 0.24 [0.21; 0.27] | 9.3% |
| Random effects model | | | | 3804 | 0.19 [0.11; 0.29] | 18.7% |
| Heterogeneity: $I^2 = 98\%$, $\tau^2 = 0.0073$, $p < 0.01$ | | | | | | |
| United States | | | | | | |
| Cohen 2005 | 03/99-02/00 | 0-59 | 138 | 684 | 0.20 [0.17; 0.23] | 9.3% |
| Mast 2009 | 02/05-07/06 | 0-59 | 474 | 748 | 0.63 [0.60; 0.67] | 9.3% |
| Random effects model | | | | 1432 | 0.41 [0.06; 0.82] | 18.6% |
| Heterogeneity: $I^2 = 100\%$, $\tau^2 = 0.1028$, $p < 0.01$ | | | | | | |
| **Random effects model** | | | | **7764** | **0.29 [0.20; 0.39]** | **100.0%** |
| **Prediction interval** | | | | | **[0.02; 0.70]** | |
| Heterogeneity: $I^2 = 99\%$, $\tau^2 = 0.0316$, $p < 0.01$ | | | | | | |
| Residual heterogeneity: $I^2 = 99\%$, $p < 0.01$ | | | | | | |

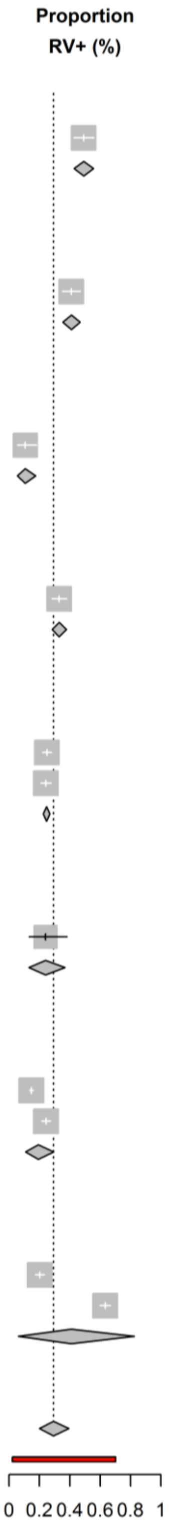


RVGE: Rotavirus gastroenteritis; RV+: stools samples that tested positive for rotavirus; AGE: acute gastroenteritis; CI: Confidence Interval

**Supplementary Figure 17.** Incidence rate of hospitalisations for RVGE of children aged 0-5 years per 100 000 person-years, including only studies with good external validity (forest plot)

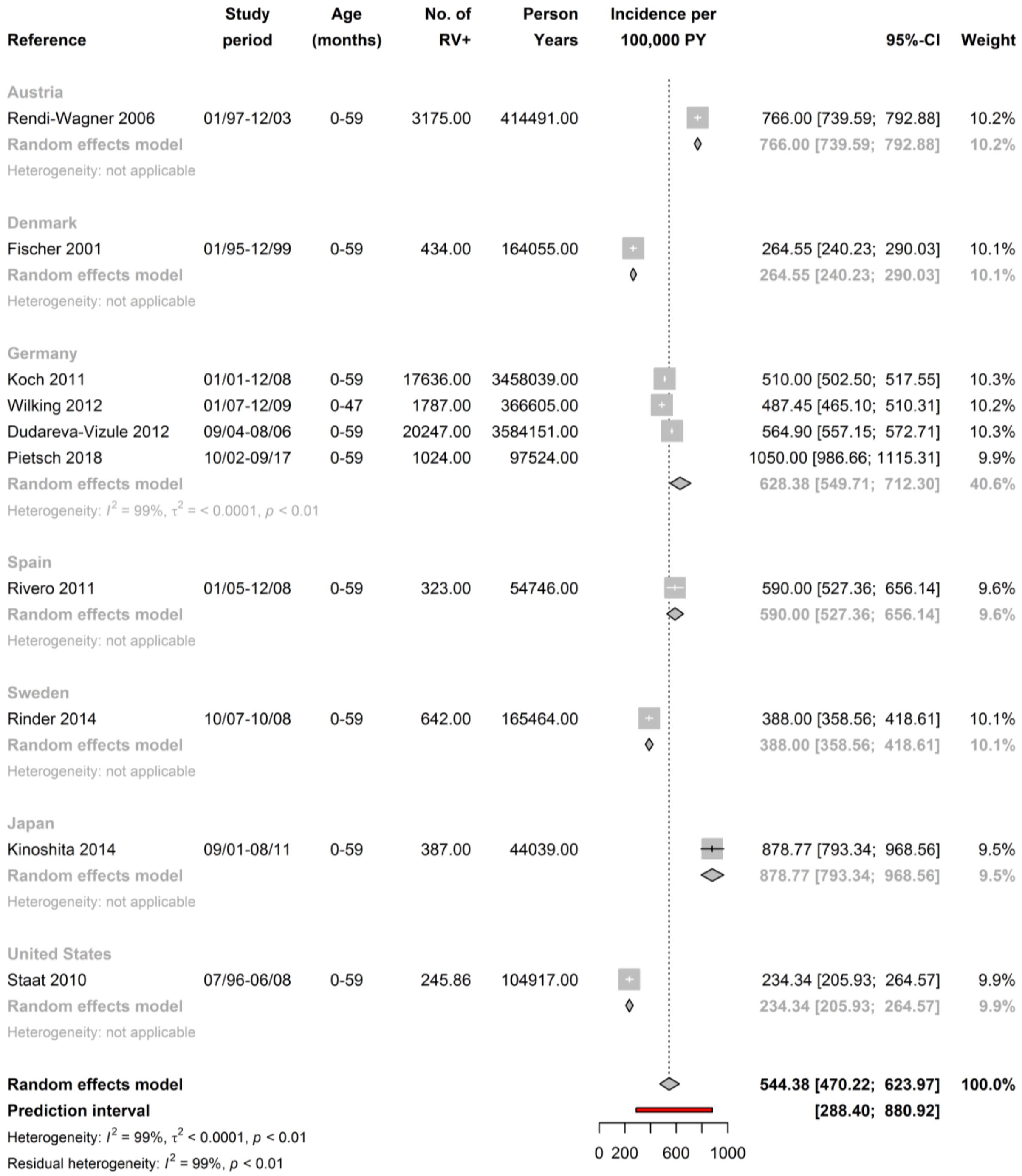


RVGE: Rotavirus gastroenteritis; RV+: stools samples that tested positive for rotavirus; PY: person-years; CI: Confidence Interval

**Supplementary Figure 18**. Proportion of hospitalisations for RVGE among all hospitalisations for acute gastroenteritis of children aged 0-5 years, including only studies with good external validity (forest plot)

| Reference | Study period | Age (months) | No. of RV+ | Total AGE | Proportion RV+ (%) | 95%-CI | Weight |
|---|---|---|---|---|---|---|---|
| **Belgium** | | | | | | | |
| Raes 2007 | 06/02-05/06 | 0-59 | 423 | 1074 | | 0.39 [0.36; 0.42] | 3.3% |
| Raes 2011 | 06/04-05/06 | 0-59 | 881 | 3161 | | 0.28 [0.26; 0.29] | 3.4% |
| Random effects model | | | | 4235 | | 0.33 [0.23; 0.45] | 6.7% |
| Heterogeneity: $I^2 = 98\%$, $\tau^2 = 0.0073$, $p < 0.01$ | | | | | | | |
| **Czech Republic** | | | | | | | |
| Arientova 2012 | 09/06-12/08 | 0-59 | 478 | 832 | | 0.57 [0.54; 0.61] | 3.3% |
| Random effects model | | | | 832 | | 0.57 [0.54; 0.61] | 3.3% |
| Heterogeneity: not applicable | | | | | | | |
| **Denmark** | | | | | | | |
| Fischer 2001 | 01/95-12/99 | 0-59 | 434 | 2146 | | 0.20 [0.19; 0.22] | 3.4% |
| Fischer 2011 | 02/09-03/10 | 0-59 | 1074 | 2610 | | 0.41 [0.39; 0.43] | 3.4% |
| Random effects model | | | | 4756 | | 0.30 [0.12; 0.52] | 6.7% |
| Heterogeneity: $I^2 = 100\%$, $\tau^2 = 0.0263$, $p < 0.01$ | | | | | | | |
| **France** | | | | | | | |
| Forster 2009 | 02/05-08/06 | 0-59 | 186 | 289 | | 0.64 [0.59; 0.70] | 3.3% |
| Random effects model | | | | 289 | | 0.64 [0.59; 0.70] | 3.3% |
| Heterogeneity: not applicable | | | | | | | |
| **Germany** | | | | | | | |
| Poppe 2002 | 06/97-05/98 | 0-48 | 621 | 1451 | | 0.43 [0.40; 0.45] | 3.4% |
| Forster 2009 | 02/05-08/06 | 0-59 | 380 | 621 | | 0.61 [0.57; 0.65] | 3.3% |
| Koch 2011 | 01/01-12/08 | 0-59 | 17636 | 25154 | | 0.70 [0.70; 0.71] | 3.4% |
| Pietsch 2018 | 10/02-09/17 | 0-59 | 1024 | 3226 | | 0.32 [0.30; 0.33] | 3.4% |
| Random effects model | | | | 30452 | | 0.52 [0.29; 0.74] | 13.4% |
| Heterogeneity: $I^2 = 100\%$, $\tau^2 = 0.0561$, $p = 0$ | | | | | | | |
| **Greece** | | | | | | | |
| Levidioutou 2009 | 01/00-12/06 | 0-59 | 904 | 4604 | | 0.20 [0.18; 0.21] | 3.4% |
| Nika 2010 | 01/08-12/09 | 0-59 | 216 | 1352 | | 0.16 [0.14; 0.18] | 3.4% |
| Konstantopoulos 2013 | 07/08-03/10 | 0-59 | 109 | 458 | | 0.24 [0.20; 0.28] | 3.3% |
| Random effects model | | | | 6414 | | 0.19 [0.16; 0.23] | 10.0% |
| Heterogeneity: $I^2 = 88\%$, $\tau^2 = 0.0013$, $p < 0.01$ | | | | | | | |
| **Italy** | | | | | | | |
| Albano 2006 | NA-NA | 1-48 | 381 | 911 | | 0.42 [0.39; 0.45] | 3.3% |
| De Grazia 2006 | 01/01-12/05 | 0-59 | 411 | 1030 | | 0.40 [0.37; 0.43] | 3.3% |
| Forster 2009 | 02/05-08/06 | 0-59 | 66 | 199 | | 0.33 [0.27; 0.40] | 3.3% |
| De Grazia 2014 | 01/85-08/12 | 0-59 | 2138 | 6522 | | 0.33 [0.32; 0.34] | 3.4% |
| Random effects model | | | | 8662 | | 0.37 [0.32; 0.42] | 13.3% |
| Heterogeneity: $I^2 = 93\%$, $\tau^2 = 0.0028$, $p < 0.01$ | | | | | | | |
| **Spain** | | | | | | | |
| Sanchez-Fauquier 2003 | 10/98-09/02 | 0-59 | 1155 | 3760 | | 0.31 [0.29; 0.32] | 3.4% |
| Forster 2009 | 02/05-08/06 | 0-59 | 86 | 164 | | 0.52 [0.45; 0.60] | 3.3% |
| Cilla 2010 | 07/96-06/08 | 1-59 | 419 | 1070 | | 0.39 [0.36; 0.42] | 3.3% |
| Rivero 2011 | 01/05-12/08 | 0-59 | 323 | 663 | | 0.49 [0.45; 0.53] | 3.3% |
| Gonzalez Galan 2011 | 04/06-04/07 | 0-59 | 80 | 399 | | 0.20 [0.16; 0.24] | 3.3% |
| Random effects model | | | | 6056 | | 0.38 [0.29; 0.47] | 16.6% |
| Heterogeneity: $I^2 = 97\%$, $\tau^2 = 0.0109$, $p < 0.01$ | | | | | | | |

*Supplementary Figure 18 (continued)*

| Reference | Study period | Age (months) | No. of RV+ | Total AGE | Proportion RV+ (%) | 95%-CI | Weight |
|---|---|---|---|---|---|---|---|
| **Sweden** | | | | | | | |
| Rinder 2014 | 10/07-10/08 | 0-59 | 642 | 1549 | | 0.41 [0.39; 0.44] | 3.4% |
| **Random effects model** | | | | **1549** | | **0.41 [0.39; 0.44]** | **3.4%** |
| Heterogeneity: not applicable | | | | | | | |
| **United Kingdom** | | | | | | | |
| Forster 2009 | 02/05-08/06 | 0-59 | 27 | 52 | | 0.52 [0.38; 0.66] | 3.0% |
| **Random effects model** | | | | **52** | | **0.52 [0.38; 0.65]** | **3.0%** |
| Heterogeneity: not applicable | | | | | | | |
| **Australia** | | | | | | | |
| Galati 2006 | 07/98-06/03 | 0-59 | 2344 | 4688 | | 0.50 [0.49; 0.51] | 3.4% |
| Campbell 2009 | 07/01-06/06 | 0-59 | 310 | 881 | | 0.35 [0.32; 0.38] | 3.3% |
| Jayasinghe 2012 | 01/00-12/07 | 0-59 | 689 | 1453 | | 0.47 [0.45; 0.50] | 3.4% |
| **Random effects model** | | | | **7022** | | **0.44 [0.37; 0.52]** | **10.1%** |
| Heterogeneity: $I^2$ = 97%, $\tau^2$ = 0.0047, $p$ < 0.01 | | | | | | | |
| **Japan** | | | | | | | |
| Kinoshita 2014 | 09/01-08/11 | 0-59 | 387 | 834 | | 0.46 [0.43; 0.50] | 3.3% |
| **Random effects model** | | | | **834** | | **0.46 [0.43; 0.50]** | **3.3%** |
| Heterogeneity: not applicable | | | | | | | |
| **United States** | | | | | | | |
| Lee 2005 | 11/97-12/99 | 0-59 | 548 | 2067 | | 0.27 [0.25; 0.28] | 3.4% |
| Mast 2009 | 02/05-07/06 | 0-59 | 201 | 411 | | 0.49 [0.44; 0.54] | 3.3% |
| **Random effects model** | | | | **2478** | | **0.37 [0.17; 0.60]** | **6.7%** |
| Heterogeneity: $I^2$ = 99%, $\tau^2$ = 0.0269, $p$ < 0.01 | | | | | | | |
| **Random effects model** | | | | **73631** | | **0.40 [0.32; 0.47]** | **100.0%** |
| **Prediction interval** | | | | | | **[0.05; 0.81]** | |
| Heterogeneity: $I^2$ = 100%, $\tau^2$ = 0.0451, $p$ = 0 | | | | | | | |
| Residual heterogeneity: $I^2$ = 99%, $p$ = 0 | | | | | | | |

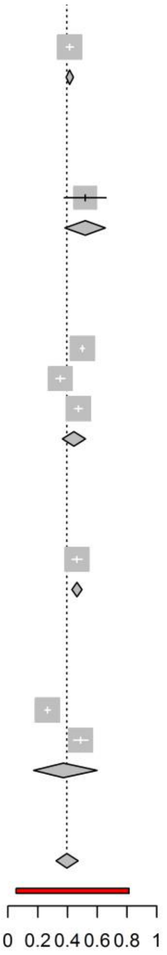


RVGE: Rotavirus gastroenteritis; RV+: stools samples that tested positive for rotavirus; AGE: acute gastroenteritis; CI: Confidence Interval

**Supplementary Figure 19**. Incidence rate of RVGE nosocomial infections among children aged 0-5 years hospitalised for reasons other than acute gastroenteritis per 100 000 person-years, including only studies with good external validity (forest plot)

| Reference | Study period | Age (months) | No. of RV+ | Person Years | Incidence per 100,000 PY | 95%-CI | Weight |
|---|---|---|---|---|---|---|---|
| Germany | | | | | | | |
| Dudareva-Vizule 2012 | 09/04-08/06 | 0-59 | 1601 | 3584151.00 | | 44.67 [42.51; 46.88] | 92.9% |
| Random effects model | | | | | | 44.67 [42.51; 46.88] | 92.9% |
| Heterogeneity: not applicable | | | | | | | |
| Greece | | | | | | | |
| Konstantopoulos 2013 | 07/08-03/10 | 0-59 | 8 | 26667.00 | | 30.00 [12.22; 55.01] | 7.1% |
| Random effects model | | | | | | 30.00 [12.22; 55.01] | 7.1% |
| Heterogeneity: not applicable | | | | | | | |
| **Random effects model** | | | | | | **42.76 [36.12; 49.96]** | **100.0%** |
| Heterogeneity: $I^2$ = 13%, $\tau^2$ < 0.0001, $p$ = 0.28 | | | | | | | |
| Residual heterogeneity: $I^2$ = NA%, $p$ = NA | | | | | | | |

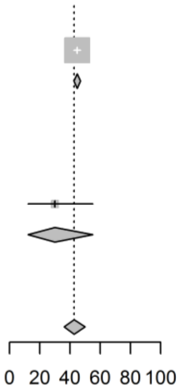


RVGE: Rotavirus gastroenteritis; RV+: stools samples that tested positive for rotavirus; PY: person-years; CI: Confidence Interval

**Supplementary Figure 21**. Incidence rate of RVGE nosocomial infections among children aged 0-5 years hospitalised for reasons other than acute gastroenteritis per 1000 hospital-days, including only studies with good external validity (forest plot)

| Reference | Study period | Age (months) | No. of RV+ | Hospital-Days | Incidence per 1,000 Hospital-Days | 95%-CI | Weight |
|---|---|---|---|---|---|---|---|
| Europe | | | | | | | |
| Forster 2009 | 02/05-08/06 | 0-59 | 117.000 | 254348.00 | | 0.46 [0.38; 0.55] | 50.1% |
| Random effects model | | | | | | 0.46 [0.38; 0.55] | 50.1% |
| Heterogeneity: not applicable | | | | | | | |
| Canada | | | | | | | |
| Verhagen 2011 | 04/98-03/08 | 0-59 | 181.282 | 229471.00 | | 0.79 [0.68; 0.91] | 49.9% |
| Random effects model | | | | | | 0.79 [0.68; 0.91] | 49.9% |
| Heterogeneity: not applicable | | | | | | | |
| **Random effects model** | | | | | | **0.61 [0.33; 0.98]** | **100.0%** |
| Heterogeneity: $I^2$ = 95%, $\tau^2$ < 0.0001, $p$ < 0.01 | | | | | | | |
| Residual heterogeneity: $I^2$ = NA%, $p$ = NA | | | | | | | |

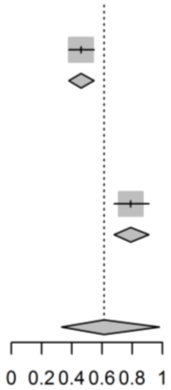


RVGE: Rotavirus gastroenteritis; RV+: stools samples that tested positive for rotavirus; CI: Confidence Interval

**Supplementary Figure 22**. Proportion of acute gastroenteritis nosocomial infections caused by RV among children aged 0-5 years hospitalised for reasons other than acute gastroenteritis, including only studies with good external validity (forest plot)

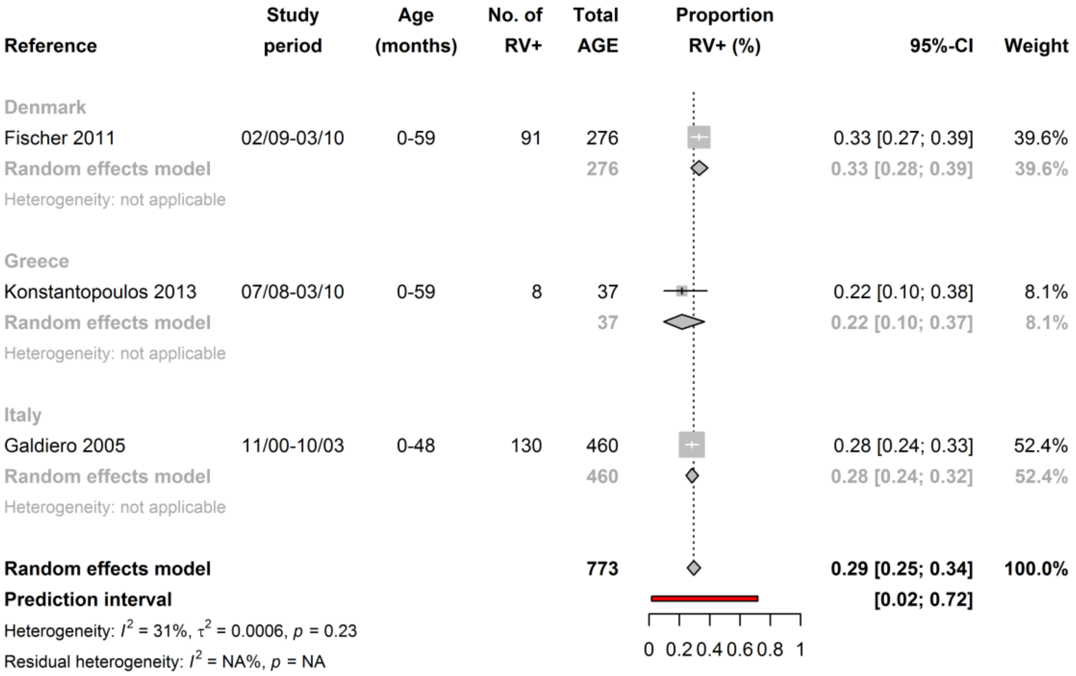


RVGE: Rotavirus gastroenteritis; RV+: stools samples that tested positive for rotavirus; AGE: acute gastroenteritis; CI: Confidence Interval
